# Data Assimilation Substitutes for Biological Complexity in Hybrid Influenza Forecasting Models

**DOI:** 10.64898/2026.05.19.26353597

**Authors:** Tijs W. Alleman, Tim Van Wesemael, Neha Shanker, Matthew S. Mietchen, Sara Loo, Oluwafeyisore Ajagbe, Jan M. Baetens, Joseph Lemaitre, Alison L. Hill, Shaun A. Truelove, Ana I. Bento

## Abstract

Infectious-disease forecasts increasingly inform public-health preparedness, yet it is unclear whether hybrid mechanistic-statistical models gain more accuracy from added biological complexity or from assimilating richer data, a choice that affects how forecasting programs invest limited resources. Using eight retrospective influenza seasons in North Carolina, we evaluate whether training on historical data and assimilating auxiliary emergency department (ED) visit data improves four-week-ahead hospital admission forecasts more than adding biological complexity (multi-subtype structure and cross-season immunity). Hierarchical Bayesian training on historical data improves accuracy by 22.4% (95% CI: 16.4–28.1%), and inclusion of ED visit data yields a further 5.3% (95% CI: 3.0–7.6%) improvement, whereas added biological complexity produces diminishing or null gains. We further observe a substitution effect in which ED visit data partially compensates for omitted biological structure. We deployed a simplified model variant in the 2025–2026 CDC FluSight Challenge and ranked among the top ensemble performers, supporting the robustness of Bayesian hierarchical training in real time. Together, we find that short-term forecast accuracy is driven more by historical learning and assimilating auxiliary signals than by biological fidelity.

## 1 Introduction

Accurate forecasts of hospital admissions are important for healthcare preparedness during the respiratory season, yet remain challenging because epidemic timing, peak magnitude, shape, and age-specific burden vary substantially across years. Seasonal influenza exemplifies this challenge, imposing a persistent global burden with approximately one billion cases and considerable hospitalizations annually [1–3], while exhibiting marked variability across seasons.

This variability arises from interacting drivers across multiple scales. Between-season factors such as antigenic drift, vaccine uptake and efficacy, and prior infection history shape initial population susceptibility [4–7]. Within-season processes, including co-circulation of types, subtypes, and lineages, waning immunity, climate and weather, and behavioral and reporting shifts, further modulate epidemic trajectories, inducing abrupt changes in reported disease burden [7–33] (Supplementary Section A.1). These dynamics are evident in the hospital admissions for influenza in North Carolina (NC) from 2014–2025 (Fig. A1) and pose fundamental challenges for short-term forecasting [32].

Traditional mechanistic models such as the SIR model provide interpretability but are prone to structural misspecification when biological complexity is incompletely represented. Conversely, statistical approaches adapt flexibly to recent trends yet sacrifice interpretability for longer-term forecasts or scenarios and may be brittle under sparse or noisy data. Hybrid mechanistic– statistical models seek to reconcile these limitations by embedding a mechanistic transmission core in a flexible statistical framework to absorb model inadequacy. Examples include particle-filtering data assimilation [34, 35], artificial-intelligence methods such as neural differential equations [36, 37], and Bayesian partial pooling [38, 39]. Large multi-model assessments have shown that ensembles of relatively simple models, when driven by standardized surveillance data, perform competitively [40–42], and recent work indicates that short-term forecastability is influenced strongly by the data-generating process [43, 44].

A central question therefore arises: should hybrid models prioritize biological complexity, for example multi-subtype or lineage resolution and inter-season cross-immunity, or instead assimilate richer data streams, such as historical data and auxiliary syndromic signals like ED visits or wastewater surveillance, to improve short-term hospital admission forecasts?

This tension reflects a classical bias-variance trade-off. Increasing biological complexity can reduce structural bias when mechanisms are well specified, but often inflates estimation variance through a larger parameter space, especially when data are limited. In contrast, training on historical data and incorporating auxiliary observations of the same underlying process can constrain the latent epidemic trajectory without expanding model dimensionality, potentially reducing both bias and variance. This trade-off is not unique to influenza but arises broadly in forecasting problems where mechanistic structure competes with data-driven adaptability.

The answer has direct implications for public-health agencies, informing whether limited resources are better spent on developing increasingly complex mechanistic models or in collecting, curating, and assimilating additional covariate data. To our knowledge, this trade-off has not been systematically isolated and evaluated within a unified forecasting framework.

Here, we embed a multi-strain Susceptible–Infected–Recovered (SIR) model with a time-varying transmission coefficient in a Bayesian hierarchical inference framework, in which season-specific SIR model parameters are modeled as exchangeable draws from a shared across-season hyperdistribution inferred from historical data [38] (Fig. 5). We then expand the mechanistic components of the model (subtype and lineage resolution, cross-immunity between seasons) and assimilate historical training data and auxiliary influenza emergency department (ED) syndromic surveillance data. We then retrospectively evaluate their impact on forecast accuracy via leave-one-out cross-validation (LOOCV) and training-history experiments on eight influenza seasons in NC (2014–2025, excluding pandemic years). We further prospectively deploy a simplified variant that uses a single-strain formulation and hospital admission data but no ED visits data in CDC’s FluSight 2025–2026 FluSight Challenge (52 U.S. states/territories) to assess the framework’s operational reliability.

We show that training on historical data is the dominant source of forecast skill, yielding a 22.4% (95% CI: 16.4 to 28.1%) improvement in four-week-ahead accuracy as measured using the relative Weighted Interval Score (WIS) [45], whereas assimilation of auxiliary ED visit data adds a smaller but still meaningful 5.3% (95% CI: 3.0 to 7.6%) gain. Both effects exceed those of mechanistic elaborations such as increased strain resolution and inter-season cross-immunity, which produced diminishing and null improvements. The inclusion of ED visits also reduced the benefits of additional strain granularity, consistent with a substitution effect in which an auxiliary data stream compensates for omitted biological structure. As the training dataset expands, accuracy improves with diminishing returns, and the magnitude of total improvement varies across seasons. Finally, our prospective 2025–2026 CDC FluSight entry ranked at the top of the ensemble on multiple metrics (as of March 21, 2026), supporting the robustness of the Bayesian hierarchical training approach in real time. Together, our results indicate that curating historical surveillance and maintaining auxiliary data streams offer a more dependable route to improving forecast skill than increasing biological complexity, highlighting that investments in surveillance data may provide greater gains than investments in increasingly detailed mechanistic models.

## 2 Results

### 2.1 Influence of model configuration on accuracy

Training on the eight available influenza seasons for NC yielded a 22.4% (95% CI: 16.4–28.1%) reduction in geometric mean relative WIS compared with historically uninformed models (Table 1, A3). This substantial improvement was consistent across all model configurations and baseline models, indicating that the Bayesian hierarchical training procedure effectively leverages historical data to improve four-week ahead forecast accuracy. Based on this result, all subsequent analyses were restricted to historically informed models.

**Table 1.**
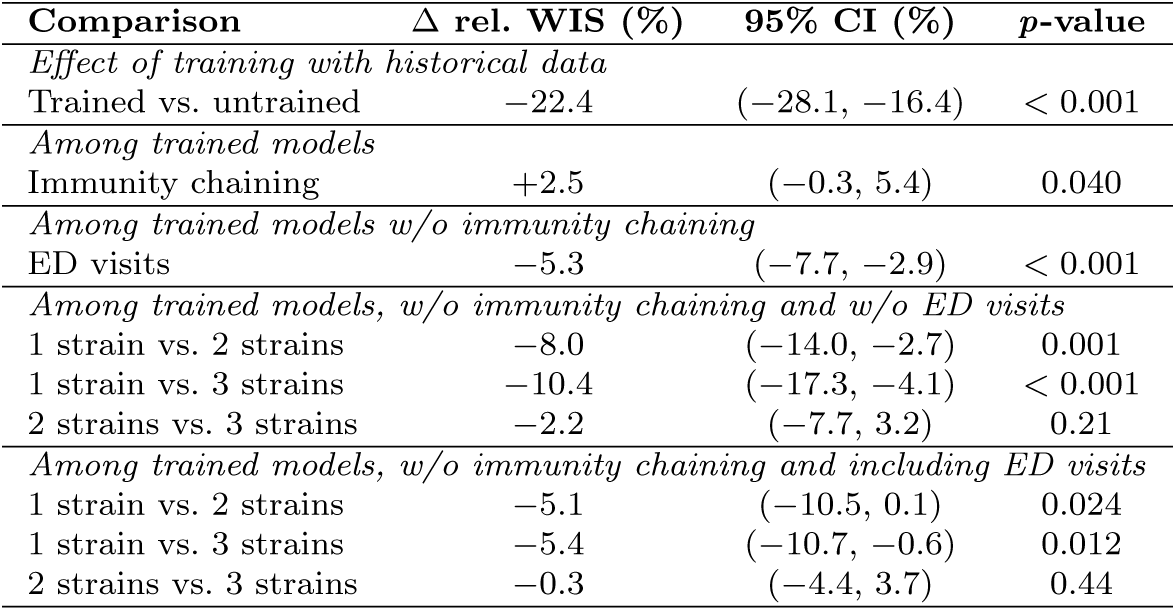
Accuracy differences between twenty-four model variants across eight North Carolina influenza seasons. Negative values indicate improved accuracy. Training on historical data, the inclusion of ED visits and the inclusion of strain granularity improved accuracy, while immunity chaining worsened accuracy. The improvements caused by adding more strain granularity show diminishing gains, especially when ED visits are already included. Results obtained from the reference-date paired bootstrap analysis (Section 4.2.1). Accuracy is expressed as the geometric mean Weighted Interval Score relative to a stationary Geometric Random Walk baseline model (Section B.3, Fig. A3).

Immunity chaining (including cross-immunity between subsequent seasons) was not associated with any reliable benefit in the geometric mean relative WIS (2.5%, 95% CI: -0.3–5.4%) (Table 1), indicating no reliable benefit. Immunity chaining attenuated the gains from auxiliary ED visit data in two- and three- strain models and reduced the performance difference between two- and three- strain formulations when ED visits were excluded (Fig. 1). We now omit model variants using immunity chaining from the paired bootstrap analysis and focus our attention on the use of the auxiliary ED visit data.

**Fig. 1.**
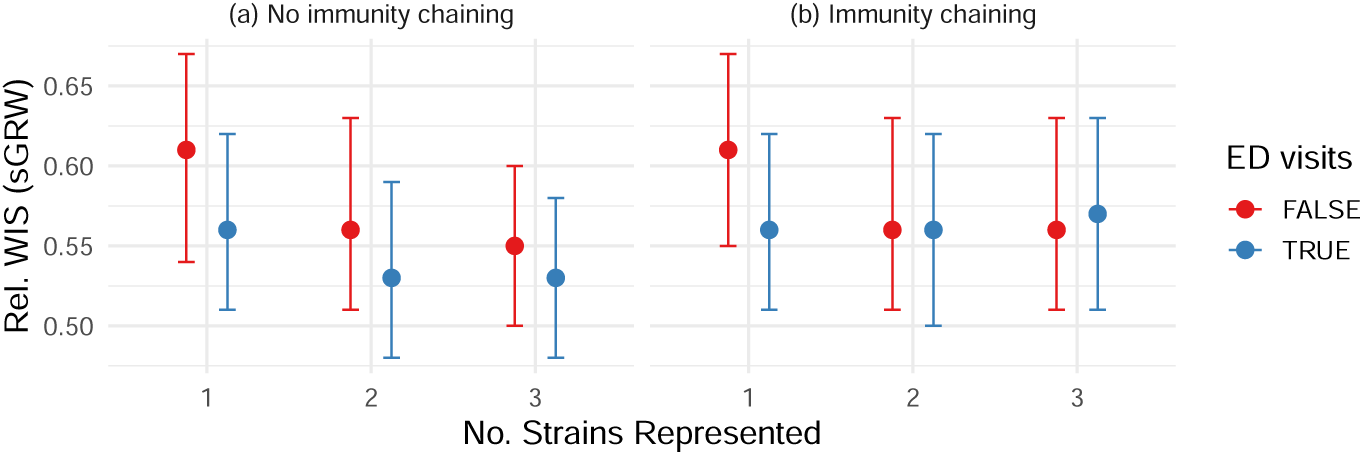
Accuracy of twelve historically informed model variants across eight North Carolina influenza seasons. Lower values correspond to more accurate forecasts. The inclusion of ED visits improved accuracy, and so did the inclusion of strain granularity, albeit with diminishing gains. Accuracy is expressed as the geometric mean Weighted Interval Score relative to a stationary Geometric Random Walk baseline model (Section B.3, Fig. A3).

Inclusion of auxiliary ED visit data during training and forecasting reduced geometric mean relative WIS by 5.3% (95% CI: 3.0–7.6%), indicating a consistent improvement in forecast accuracy (Table 1). This effect is also evident in Fig. 1, where ED visit inclusion improves performance regardless of the number of strains represented in the model. The average delay between infections and ED visits inferred during training was short, *θ* = 1.4 *d.* (95 % CI: 0.4–3.5 days), which is small relative to the weekly aggregation of the forecast target and suggests that ED visits act primarily as an additional, partially independent observation of the same infection process rather than as a strongly leading indicator.

Finally, among trained models excluding immunity chaining, multi-strain formulations consistently outperformed the single-strain model. Without auxiliary ED visit data, forecast accuracy improved monotonically from one to two to three strains, although the difference between the two strain and three strain formulations is not statistically significant. With ED visits included, both two- and three-strain models significantly outperformed the single-strain formulation, but no detectable difference was observed between the two- and three-strain models (0.3% difference; 95% CI: -3.7–4.4%) (Table 1, Fig. 1). These results indicate diminishing returns to increased strain resolution once auxiliary data are incorporated. Consistent with this pattern, no accuracy difference was detected between a single-strain model with ED visits and a two-strain model without ED visits.

The fitted coefficients of the linear mixed-effects model corroborate the paired bootstrap results regarding the importance of each model addition (Table A2). Immunity chaining showed no consistent benefit and was associated with a small increase in relative WIS, while inclusion of auxiliary ED visit data was associated with a significant improvement in forecast accuracy. Relative to the single-strain formulation, both two- and three-strain models were associated with statistically significant accuracy improvements.

The trajectories of the transmission-rate modifiers Δ*β*(*t*) in NC are shown in Figure 2. These modifiers are directly related to the effective reproduction number. A notable feature of the estimated trajectories is their pronounced seasonal structure: modifiers are typically below zero in October, rise through early winter, peak in January–February, and then decline toward seasons’ end in April.

**Fig. 2.**
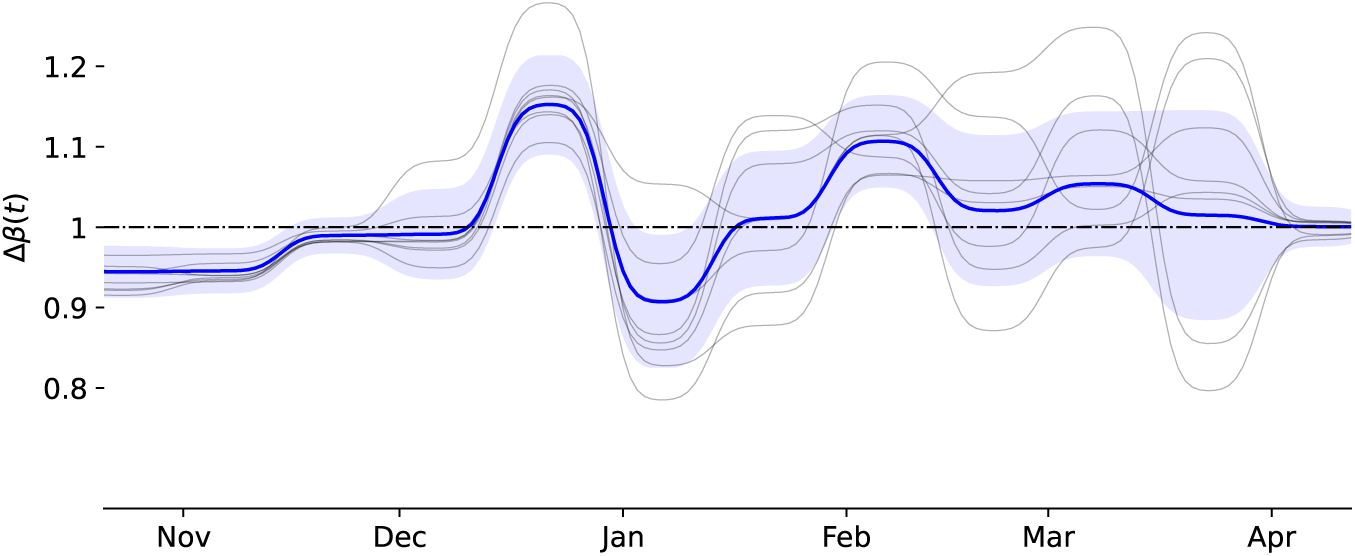
Posterior distribution of the transmission rate modifiers Δ*β*(*t*) inferred from eight North Carolina influenza seasons. Trend across the training seasons with one-sigma confidence interval (blue). Individual training seasons (grey). Modifiers show pronounced seasonal structure with a strong post-holiday reduction in transmission. For the one-strain (aggregated) model without immunity chaining and using the ED visits data (Fig. A4).

More specifically, transmissibility remains near one through mid-December, followed by a sharp increase around the holiday period, with the modifier rising from approximately 1.0 to 1.15. This surge is followed by an equally sharp decline in early January, dropping to roughly 0.9, consistent with a strong post-holiday reduction in transmission. A secondary peak occurs in early February, with modifier values near 1.1, after which the cross-season mean gradually returns to one by late April.

The grey trajectories of the season-specific modifiers indicate that uncertainty in transmissibility is greatest after the second transmission peak, consistent with late-season dynamics potentially driven by multiple circulating strains. The trajectories also exhibit clear temporal dependence, suggesting heteroskedastic autoregressive structure in Δ*β*(*t*).

### 2.2 Influence of training history on accuracy

Forecast accuracy improves with increasing training history, with clear diminishing returns; however, both the magnitude of improvement and the rate at which it is achieved vary substantially across seasons and model configurations (Fig. 3). The fitted nonlinear mixed-effects learning model quantifies these learning dynamics and decomposes them into asymptotic performance (*µ*), total accuracy gains from training (Δ), and learning rates (*κ*) (Eq. (7), Table A4).

**Fig. 3.**
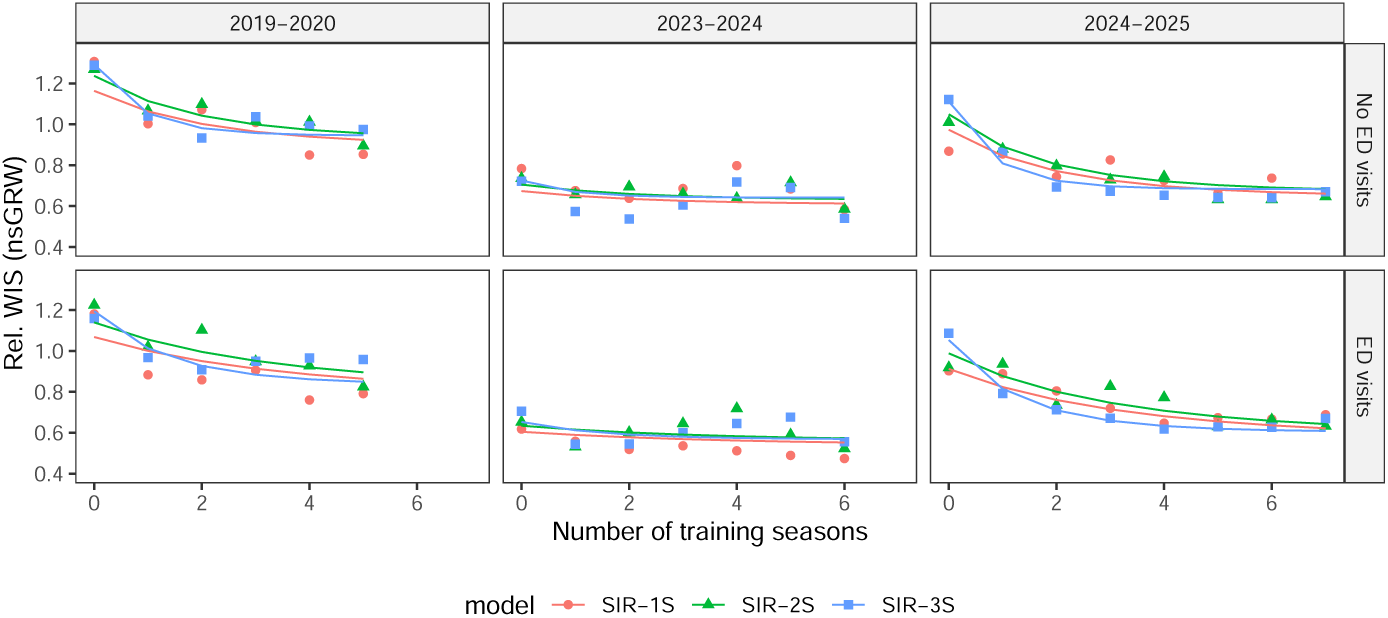
Forecast accuracy in the 2019–2020 (left), 2023–2024 (middle) and 2024– 2025 (right) North Carolina influenza seasons as a function of the number of preceding seasons used for training. Lower values correspond to more accurate forecasts. Seasonal context is the dominant determinant of how much training improves performance. Accuracy is expressed as the geometric mean Weighted Interval Score relative to the non-stationary Geometric Random Walk baseline model (Section B.3, Fig. A7). Markers represent the empirical accuracy, while full lines represent the modeled accuracy (Eqs.(B14), (B15)). Models do not use immunity chaining.

Given a sufficiently long training history, all model configurations outperform the baseline models across the three focal seasons, as indicated by the intercept for *µ* being smaller than the mean log WIS of the baseline models (Table A4).

Increasing strain resolution yields modest, monotonic improvements in asymptotic accuracy that fail to reach statistical significance. In contrast, inclusion of ED visits leads to a consistent and statistically significant improvement in asymptotic accuracy across both baselines. These modest and diminishing gains from adding in strain detail and the stronger gains from ED visit inclusion are consistent with the results of the LOOCV analysis (Table 1).

Models trained with no historical data perform substantially worse than their asymptotic limit, reaffirming that Bayesian hierarchical training contributes meaningfully to forecast accuracy, as indicated by the negative intercept for Δ (Table 1).

Seasonal context is the dominant determinant of how much training improves performance. In the 2023–2024 season, the total accuracy gain from training is minimal, suggesting limited benefit from additional historical data. In contrast, training yields substantially larger gains in 2024–2025, indicating that historical information was particularly informative in that season. Strain resolution and ED visit inclusion have little effect on the total amount of accuracy gained, implying that their benefits primarily shift asymptotic performance rather than increasing overall learnability.

Learning rates differ systematically across model configurations. Models with greater strain resolution exhibit faster learning, while inclusion of ED visits slows the rate at which accuracy approaches its asymptotic level, despite improving ultimate performance. Thus, ED visit data introduces a clear speed–accuracy trade-off: it enhances asymptotic accuracy but requires longer training histories for these gains to fully materialize.

### 2.3 Participation in the CDC FluSight Challenge

Among the 19 models with 100 % of forecasts submitted, Cornell JHU-hierarchSIR ranked first on arithmetic mean relative MAE, third on arithmetic mean relative WIS, and tenth on geometric mean relative WIS. Our model outperformed the FluSight Ensemble on the arithmetic mean MAE and WIS but not on the geometric mean WIS. Predictive interval coverage was below nominal levels: 50 % and 95 % intervals achieved 38.4 % and 83.4 % empirical coverage, respectively. In comparison, the FluSight Ensemble achieved 51.8 % and 90.0 % coverage [46, 47]. We found the end-of-season ranking is highly sensitive to the omission of forecasts, motivating our choice to only compare across models with 100 % submission completeness (Fig. A9).

Comparing rolling accuracy to observed hospital admissions (Fig. 4A,B) shows that the model performed best when admissions were high, ranking first on arithmetic mean relative MAE and WIS. Performance on geometric mean relative WIS peaked at fourth rank in January 2026 and declined noticeably toward the end of the challenge.

**Fig. 4.**
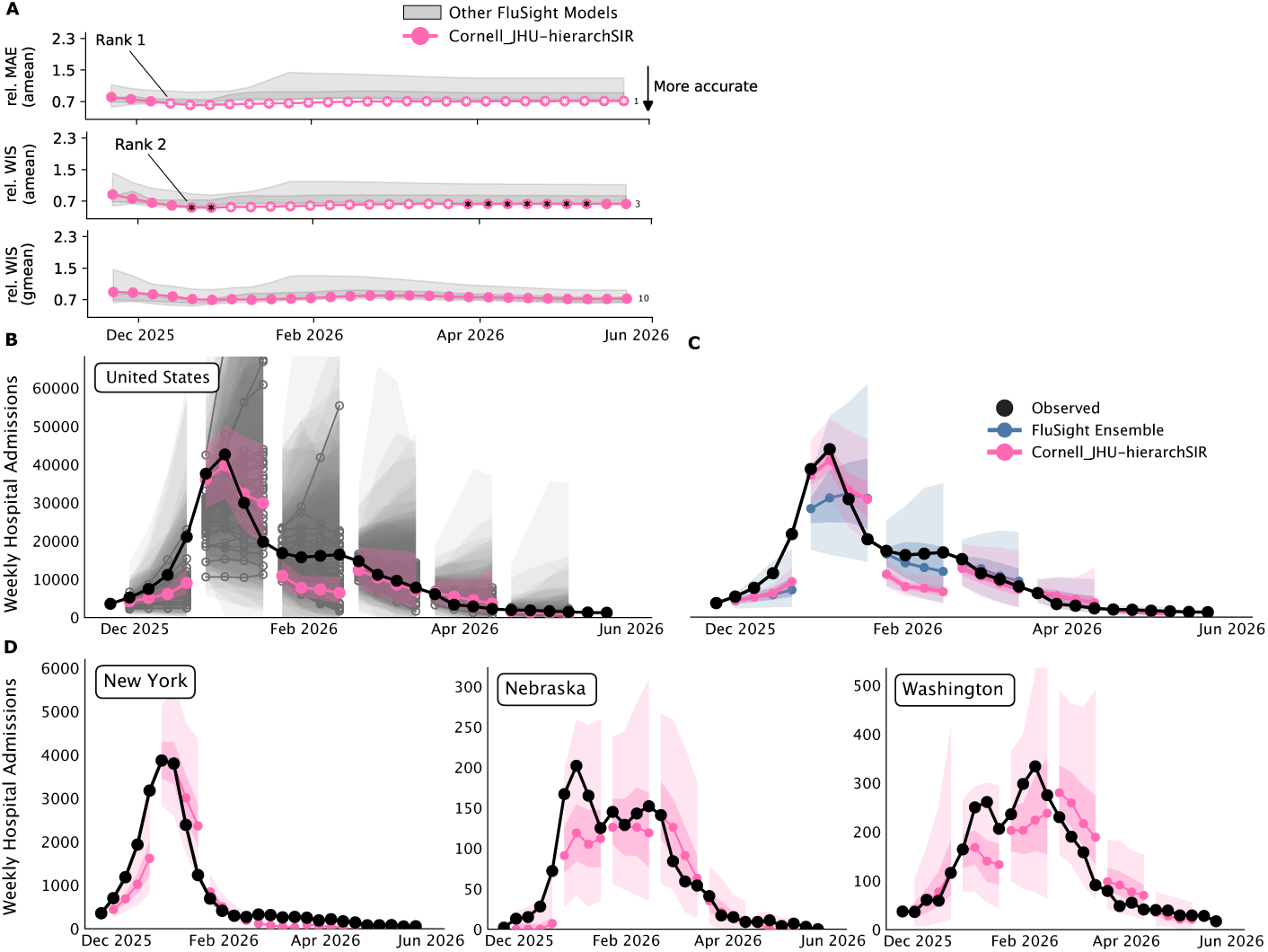
Accuracy, ranking and forecasts of our prospective participation in the 2025–2026 CDC FluSight Challenge. **(A)** Rolling accuracy and ranking of Cornell JHU-hierarchSIR versus other participating models with 100 % submission completeness. **(B)** Our forecasts for the U.S. compared to all other participating models. **(C)** Our forecasts compared to the FluSight Ensemble. **(D)** Our forecasts for New York, Nebraska, and Washington, which show influenza moving from the east to west coast during the 2025–2026 season.

Forecast trajectories generally track the direction of observed admissions at both the national and state levels, capturing major upward and downward trends and rarely diverging markedly from the FluSight Ensemble (Figure 4C,D). Performance was particularly strong in several East Coast states, where early epidemic onset caused the peak to coincide with the holiday period and its associated transmission modifier shifts (Fig. 2; New York, Maryland, and North Carolina constituted the top three on arithmetic mean relative WIS).

In contrast, performance was weaker in states with later epidemic onset and lower populations (arithmetic mean rel. WIS *>* 1 in the 10 lowest-ranked U.S. states and territories: Puerto Rico, Vermont, Alaska, Oklahoma, Michigan, Arizona, Arkansas, Wyoming, and California – in increasing order of performance; Fig. A8). Forecast intervals were notably narrower than those of the FluSight Ensemble and most participating models [47], consistent with the observed under-coverage.

## 3 Discussion

We demonstrated that a canonical SIR model with flexible transmission discrepancy, trained hierarchically on historical data and augmented with an auxiliary syndromic signal, outperforms both historically uninformed mechanistic models and more biologically elaborate models. The dominant accuracy gain came from Bayesian cross-season hierarchical training, with ED visits assimilation providing a smaller but consistent additional improvement. In contrast, finer strain resolution and inter-season immunity chaining yielded diminishing or null gains, shifting the hybrid mechanistic-statistical model toward a less favorable point on the bias-variance frontier.

This pattern aligns with a growing body of work showing that respiratory disease forecasts improve when models are able to exploit historical data and are tightly coupled to informative surveillance data [35, 38, 48]. Hybrid mechanistic-statistical frameworks that borrow strength from historical data outperform uninformed alternatives [35, 38] and large multi-model assessments such as the CDC FluSight Challenge have likewise shown that ensembles of models driven by standardized surveillance data perform competitively [40–42]. Recent work on the forecastability of infectious disease time series further indicates that near-term predictability is mostly influenced by the data-generating process and reporting delays [43, 44]. Within this landscape, our results show, in a single hybrid framework and through systematic ablations, that the marginal accuracy returns of additional biological state variables are small relative to the returns from historical training and from including a second informative and temporally leading data stream.

From our retrospective experiments in NC, we draw three design principles for short-term forecasting with hybrid mechanistic-statistical models. First, Bayesian hierarchical training on historical data is the dominant source of forecast skill. Exploiting cross-season information substantially improved relative WIS by 22.4 % (95 % CI: 16.4–28.1 %) compared to historically uninformed models, with gains plateauing after 4–5 seasons [35]. In our framework, across-season hyperdistributions capture recurrent seasonal regularities, like the holiday-period transmission signature, and act as informative priors that stabilize within-season inference, particularly early in the season when data are sparse. At the same time, learnability varied sharply across seasons, with minimal gains in 2023–2024 and steep gains in 2024–2025, which indicates that epidemic novelty bounds the usefulness of historical information. A pandemic represents an extreme case of such novelty, where historical scaffolding is uninformative.

Second, the inclusion of ED visits represents a significant but more modest secondary gain of 5.3 % (95 % CI: 2.9–7.7 %). Adding ED visit data consistently improved forecast accuracy across all model configurations, even though the inferred lead of ED visits relative to hospital admissions was short compared with the weekly aggregation of the target. We therefore interpret the improvement as arising primarily from a partially independent observation of the same underlying infection process, which narrows posterior uncertainty about the latent epidemic trajectory and the time-varying transmission coefficient. This interpretation is consistent with prior demonstrations that auxiliary indicators, including internet search, electronic health records, and ED-based syndromic surveillance, improve influenza forecasts [49–51]. Once ED visits are included, additional strain granularity yields diminishing returns, suggesting a substitution effect in which an auxiliary data stream can compensate for omitted biological structure in short-horizon hospital-admission forecasts.

Third, time-varying transmission modifiers appear to capture recurrent behavioral signatures that are exploitable for forecasting. Estimated transmission trajectories showed reproducible seasonal patterns (Fig. 2), consistent with documented holiday-period changes in contact patterns and with the broader observation that human behavior is a major and often underappreciated driver of epidemic forecast difficulty [25, 26, 29, 32]. Encoding these recurrent behavioral regularities into the transmission modifiers is likely a key component of the gains we attribute to hierarchical training. The season-specific trajectories also exhibit temporal dependence and heteroskedasticity, which suggests that future work could improve performance further by explicitly modeling the persistence and volatility of transmission.

Inter-season immunity chaining did not improve short-horizon forecast accuracy and modestly reduced the benefit of ED visit inclusion. Two explanations are plausible. First, short-term hospital admissions may be driven more by within-season processes, such as subtype cocirculation, intraseason waning, and behavioral modulation, than by accumulated cross-season immunity [4–6, 17, 19, 21, 25, 26, 29, 32] (Supplementary Section A.1). Second, the immunity-chaining mechanism may be misspecified because it omits vaccination and cross-protection, or because immunity accumulation is strongly nonlinear [4–7, 9]. Both explanations are consistent with earlier work emphasizing the limited identifiability of cross-season immunity from admissions data alone [7]. In this respect, our results complement rather than contradict the biological literature: they suggest that, for short-horizon admission forecasting, the signal from cross-season immunity is either too weak or too poorly identified to improve forecasts.

Prospective participation in the 2025–2026 CDC FluSight Challenge showed that the framework scales operationally across 52 U.S. states and territories [40–42]. A simplified model trained on only two prior seasons ranked among the top models on arithmetic mean relative WIS and relative MAE, although it ranked lower on geometric mean relative WIS, likely because uncertainty was underestimated due to the short training dataset (Fig. A8). The difference between arithmetic and geometric rankings also underscores the importance of evaluating forecasts with multiple metrics, since each emphasizes different aspects of performance [40–42, 45]. In particular, the geometric mean relative WIS downweights the importance of epidemic peaks and longer forecast horizons, which may not fully reflect the public health priorities of a forecasting challenge focused on operational utility [40]. Using the arithmetic mean of the U.S. state or territory-specific arithmetic mean relative WIS would preserve equal weighting across locations while maintaining sensitivity to forecast horizon and epidemic intensity.

This study has several limitations. The retrospective analyses used North Carolina data with detailed strain typing and auxiliary syndromic information, so generalization to other jurisdictions and pathogens remains to be tested, although the FluSight deployment suggests that the hierarchical training component scales [40, 41]. Strain independence in the SIR model assumes limited short-term competition for susceptible hosts, which is reasonable for short-horizon forecasts but may be limiting for longer-term outcomes such as peak timing [18]. The transmission discrepancy also lacks explicit autoregressive volatility structure, which is a promising direction for refinement [52]. Finally, we focused on ED visits as the auxiliary signal; other temporally informative streams, including insurance claims, weather, wastewater surveillance, and social media, could be compared systematically within the same framework [48, 50, 51].

Together, these results position the framework as operationally robust and supporting a structured approach to model biological complexity in respiratory disease forecasting. Mechanistic structure remains important for interpretability and for connecting short-term forecasts to questions about evolution, immunity, and interventions [5, 24]. For short-term hospital-admission forecasting, however, the largest gains came from historical data and auxiliary surveillance, suggesting that future improvements will likely depend first on assimilating richer data streams, and only then on adding biological detail [40, 41, 44]. For the programs that produce these forecasts, this indicates that returns to surveillance investment are likely to exceed returns to added model complexity.

## 4 Methods

### 4.1 Modeling Framework

#### 4.1.1 Forward simulating SIR model

##### Disease transmission

The transmission of influenza during a season is simulated using a strain-stratified SIR model. Although influenza viruses are formally classified into types, subtypes, and lineages, we use the abstract term “strain” from hereon, as it is consistent with the level of biological resolution needed in the model. We consider three resolutions of strains, ranging from granular to coarse: three strains, influenza A/H1N1pdm09, influenza A/H3N2, and influenza B; two strains, influenza A and influenza B; and a single-strain representation. Explicitly modeling sequential infections, cross-immunity, and competition for susceptible hosts would introduce substantial additional complexity and parameters with limited identifiability [7, 18]. We therefore model strains independently as a parsimonious approximation for short-term, within-season forecasting.

The transmission of influenza strain *i* is governed by the following temporally-forced deterministic variant of the canonical SIR model [53],

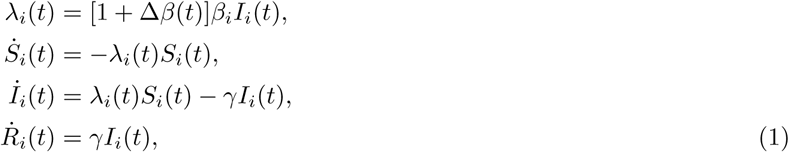

where *S_i_*(*t*) + *I_i_*(*t*) + *R_i_*(*t*) = 1. In this model, *λ_i_*(*t*) is the force of infection at day *t*, *β_i_* represents the baseline transmission coefficient of the influenza strain *i* and *γ* = 1*/*3.5 *d*^-1^ is the average duration of infectiousness, chosen to correspond to the CDC Yellow Book estimate of the most contagious phase of influenza infection [54].

##### Time-varying transmission coefficient

Δ*β*(*t*) represents a time-varying multiplicative modifier to the baseline transmission coefficient, obtained by smoothing a piecewise-continuous trajectory of twelve 15-day modifiers Δ*B_l_* with a Gaussian filter. For the influenza season of year *{*X *−* X + 1*}*, the first modifier represents the second half of October of year X and the last modifier represents the first half of April of year X + 1. Outside of this range, there is no modification of the transmission coefficient and hence Δ*β*(*t*) = 0.

This approach extends the flexibility of the SIR model, allowing it to better capture complex patterns in the observed incidence data, effectively acting as a discrepancy model in the transmission domain [38].

##### Observations

To anchor the observation’s timescale and avoid unidentifiability, we assume ED visits are contemporaneous to infections and estimate only the delay from infection to hospital admission [49]. If *I*_new*,i*_(*t*) = *λ_i_*(*t*)*S_i_*(*t*)*T* denotes the incidence of new infections for strain *i* at time *t*, with *T* equal to the total population,

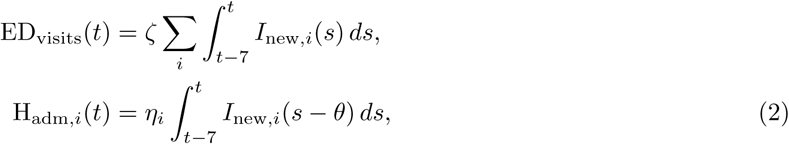

where *ζ* and *η_i_* are ascertainment probabilities for ED visits and hospital admissions respectively, and *θ* denotes the delay between infection and hospital admission.

##### Initial conditions and immunity chaining

We model seasonal influenza on a season-by-season basis. For the influenza season of year *{*X–X + 1*}*, simulations are started on October 1st of year X and run until May 1st of year X + 1. We model the initial condition of strain *i* in season *j* as follows,

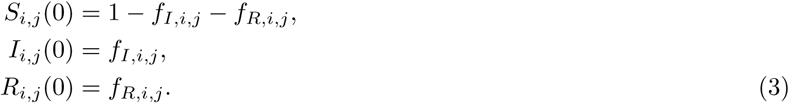

Here *f_I,i,j_* represents the initial fraction of the population infected with influenza strain *i* at the start of season *j*, and *f_R,i,j_* represents the initial fraction of the population with immunity to influenza strain *i* at the start of season *j*.

Instead of directly inferring the population’s immunity at the start of the season *f_R,i_*, we could model (within-strain) inter-season cross-immunity, i.e. the belief that the population’s immunity to influenza strain *i* at the start of a season *j* is linked to the cumulative incidence of strain *i* in prior seasons [7, 55]. Mathematically,

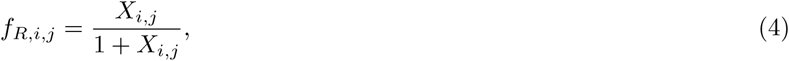

where *X_i,j_* is a linear function of the cumulative number of infections to strain *i* in the past three seasons,

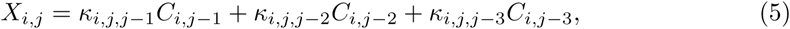

and where *C_i,j−_*_1_ represents the cumulative number of hospital admissions to strain *i* in season *j −* 1 and *κ_i,j,j−_*_1_ represents the proportion of all infections with strain *i* in season *j −* 1 that confer immunity to infection by strain *i* in season *j*. Eq. (4) is a saturating link function between past incidence and population immunity to preventing population-wide protection under heavy prior transmission. Equations (4) and (5) are from hereon referred to as *immunity chaining*. Table A1 contains a summary of the forward simulation model’s parameters.

#### 4.1.2 Bayesian Inference Framework

##### Training

We embed the forward-simulating SIR model in a Bayesian hierarchical inference framework, in which season-specific SIR model parameters are modeled as exchangeable draws from a shared across-season hyperdistribution (Fig. 5) [38]. The goal of training is to infer across-season hyperdistributions governing transmission (*β_i_*, Δ*B_l_*), observation processes (*ζ*, *η_i_*), and initial conditions (*f_I,i_* and either *f_R,i_*or the immunity-linking parameters *κ_i,j,j−n_*). To this end, we sample from the posterior distribution (Eq. (B1)) using the ensemble sampler of Goodman and Weare [56, 57].

**Fig. 5.**
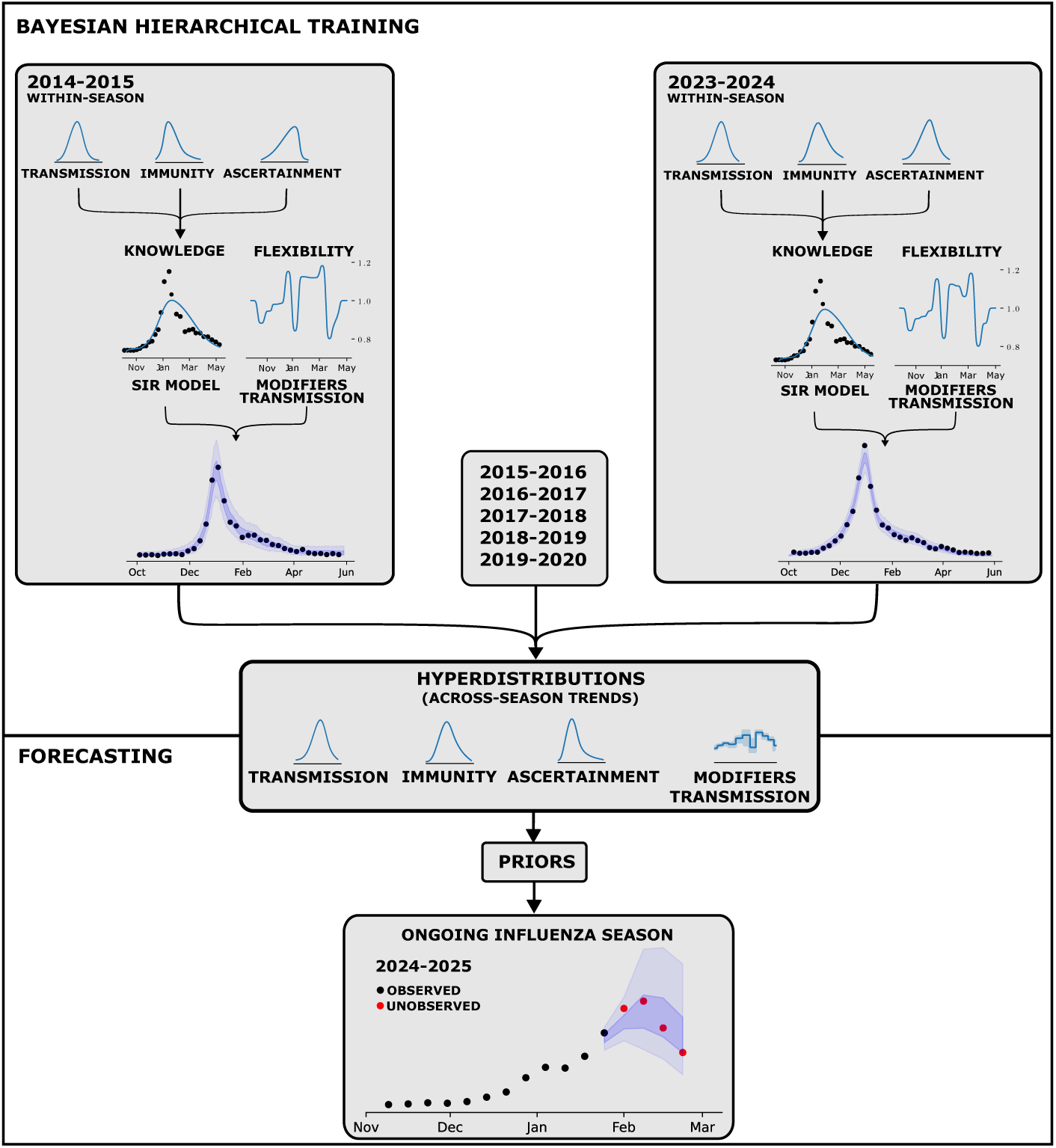
Schematic representation of the training and forecasting procedure. **<u>Training:</u>** A multi-strain SIR model with a time-varying transmission coefficient is embedded within a Bayesian hierarchical inference framework, in which season-specific SIR model parameters are modeled as exchangeable draws from a shared across-season *hyperdistribution*. The goal of training is to sample the posterior distribution of the across-season hyperdistributions. **<u>Forecasting:</u>** During an ongoing influenza season, the across-season hyperdistributions are used as informative priors for a Bayesian update of the within-season parameters. The within-season parameters are propagated through the SIR model to produce a four-week ahead predictive distribution. This two-step process allows for dynamic adaptation as new data become available.

The Bayesian inference framework simultaneously infers the within-season parameters and the parameters of the distributions that describe their variability across seasons, referred to as *cross-season hyperdistributions*. These capture cross-seasonal trends and act as priors for the within-season parameters. Details are provided in Supplementary Section B.2.

### Forecasting

The cross-season hyperdistributions are used as prior distributions for the inference of the forward simulation model’s within-season parameters during the ongoing influenza season (Fig. 5). This results in posterior distributions of the within-season parameters conditioned on the new data, which are propagated through the forward simulation model to generate a forecast distribution.

### 4.2 Forecast experiments

#### 4.2.1 Influence of model configuration on accuracy

We perform a leave-one-out cross-validation (LOOCV) across eight influenza seasons in NC (Supplementary Section B.4, Figs. A10, A11) to assess how model and training configuration affect four-week-ahead forecast accuracy on the held out season from November 15 through April 7. We evaluate 24 variants *M* obtained by varying: (i) If the model was trained to historical data, (ii) the amount of strains represented (1 strain; 2 strains: A/B; 3 strains: H1N1pdm09/H3N2/B), (iii) the inclusion of auxiliary ED visit data, and (iv) the use of immunity chaining (Eqs. (4), (5)),

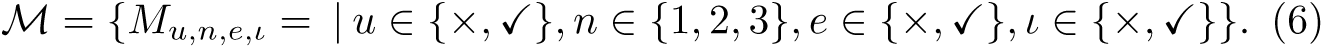

Here, *u* denotes the use of uninformative priors, reflecting a situation where the model is fit independently to each season (e.g., without borrowing strength from past seasons), *n* denotes the number of strains, *e* indicates inclusion of ED visits, and *ι* indicates the inclusion of immunity chaining.

To assess whether meaningful accuracy differences between model configurations exist, we use two complementary approaches. First, we conduct paired nonparametric bootstrap comparisons of geometric mean relative WIS [41, 42, 45, 58] (Supplementary Section B.3), pairing forecasts by forecast start date (“reference date”) to control for shared epidemic context. Second, we fit a linear mixed-effects model to the natural logarithm of the absolute WIS with a random intercept for reference date (Supplementary Section B.5).

#### 4.2.2 Influence of training history on accuracy

Focusing on the three most recent influenza seasons (2019–2020, 2023–2024 and 2024–2025), we vary the amount of historical data used for training from none (uninformative priors) up to the full available record (back to 2014– 2015). After training, we generate four-week-ahead forecasts during the focal season and compute accuracy relative to the GRW baselines (Section B.3). To characterize how forecast accuracy changes with increasing training history, we model the natural log of the absolute WIS as a nonlinear function of training length using the following mixed-effects model,

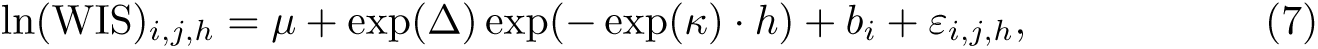

where *i* is the forecast start date (“reference date”), *j* the forecast horizon, and

*h* the number of historical seasons used to train the model. This model allows us to estimate asymptotic performance (*µ*), total accuracy gained from training (Δ), and learning rates (*κ*) while accounting for shared forecast difficulty across reference dates using mixed effects *b_i_* (Supplementary Section B.6).

#### 4.2.3 Participation in the CDC FluSight Challenge

To evaluate operational reliability and real-time forecast performance, we participated in the 2025–2026 CDC FluSight Challenge [46, 47] with the simplest viable variant of our framework named Cornell JHU-hierarchSIR.

Due to limitations in the availability of data, we restricted training to the 2023–2024 and 2024–2025 influenza seasons, modeled U.S. states and territories independently, employed a single-strain formulation, and used no ED visit data. We corrected for systematic underreporting using a generalized Dirichlet—multinomial reporting model formulated as a sequential beta-binomial survival process with coefficients estimated on a four-week rolling basis [59–61] and posterior means expressed in closed form through prior conjugacy (Supplementary Section B.7).

Each Wednesday from mid-November 2025 through the end of May 2026, we submitted a four-week ahead forecast of influenza hospital admissions in the U.S. (national), 50 U.S. states, D.C., and Puerto Rico [62]. In total, 27 forecasts across 53 locations were due during the 2025–2026 season. Forecasts were evaluated on a rolling basis using the arithmetic and geometric mean relative WIS against a stationary GRW baseline model (Supplementary section B.3, Fig. A2), as well as 50 % and 95 % forecast interval coverage, and the arithmetic relative Mean Absolute Error (MAE).

## Data Availability

The model used for North Carolina can be found in the twallema/JHU-Cornell_hierarchSIR Github repository, while the operational CDC FluSight model can be found in the BentoLab-DiseaseDynamics/Cornell_JHU-hierarchSIR repository. Detailed NC DETECT emergency department visit data are available through a Data Use Agreement with the NC Division of Public Health. More information is available at https://ncdetect.org/data-requests-for-applied-public-health-research/.

## Supplementary information

This work contains supplementary literature (Section A), methods (Section B), results (Section C) and a visualisation of the North Carolina hospital admission dataset (Section D). The model used for North Carolina can be found in the twallema/JHU-Cornell hierarchSIR Github repository, while the operational CDC FluSight model can be found in the BentoLab-DiseaseDynamics/Cornell JHU-hierarchSIR repository. Detailed NC DETECT emergency department visit data are available through a Data Use Agreement with the NC Division of Public Health. More information is available at https://ncdetect.org/data-requests-for-applied-public-health-research/.

## Author contributions

**Tijs W. Alleman**: Conceptualisation, Methodology, Software, Formal analysis, Investigation, Data curation, Writing - Original Draft, Writing - Review & Editing, Visualisation. **Tim Van Wesemael**: Software, Validation, Writing - Review & Editing. **Neha Shanker**: Conceptualisation, Data curation, Writing - Review & Editing. **Matthew Mietchen**: Conceptualisation, Data curation. **Sara Loo**: Conceptualisation, Writing - Review & Editing. **Sore Ajagbe**: Visualisation. **Jan M. Baetens**: Funding acquisition. **Joseph Lemaitre**: Conceptualisation, Writing - Review & Editing. **Alison L. Hill**: Conceptualisation, Supervision, Writing - Review & Editing, Funding acquisition. **Shaun Truelove**: Conceptualisation, Computational Resources, Supervision, Project administration, Funding acquisition. **Ana I. Bento**: Conceptualisation, Methodology, Writing - Original draft, Writing - Review & Editing, Supervision, Project administration, Funding acquisition.

## Acknowledgments

This work was supported by the Atlantic Coast Center for Infectious Disease Dynamics and Analytics (CDC CFA grant NU38FT000012), the MIDAS Coordination Center (NIGMS grant R24GM153920) and the Flemish Research Fund (grant G0G0122N). The over half million core-hours needed to perform this research were generously provided by Dr. Shaun Truelove and hosted by the Advanced Research Computing at Hopkins (ARCH) Rockfish cluster. We kindly acknowledge the work of Dr. Rebecca K. Borchering, Annabella G. Hines, Sarabeth M. Mathis, and Dr. Matthew Biggerstaff at the CDC Influenza Division for hosting the 2025– 2026 FluSight Challenge and evaluating our model’s forecasts, along with all of the challenge participants. We further acknowledge the work of Dr. Amy Ising from the University of North Carolina at Chapel Hill for her work on curating the NC DETECT data and the NC DETECT Respiratory Dashboard.

### Appendix A Supplementary literature

#### A.1 The variability between influenza seasons

The pronounced variability in seasonal influenza epidemics timing, height, shape and age-specific impact arises from the interplay of multiple processes operating across different temporal scales.

At the between-season level, antigenic drift, interannual differences in vaccine uptake and effectiveness [8, 9], and the population’s accumulated infection history [6, 7, 10–12] jointly shape susceptibility at the start of each influenza season. These factors influence not only overall epidemic size but also which influenza types, subtypes and lineages dominate or co-circulate. This composition may affect symptom severity, and consequently case ascertainment, as well as transmission characteristics such as the role of asymptomatic infections, although evidence on these effects remains mixed [13–15]. In addition, patterns of global circulation, together with global and local mobility, influence both the timing and location of viral introductions, further contributing to interseasonal heterogeneity [15, 16].

Superimposed on these between-season drivers are processes acting within individual seasons. Epidemic trajectories are shaped by the co-circulation of multiple types, subtypes and lineages of influenza [17], the location and time of their introductions [28, 33], inference with common cold viruses [18], waning of vaccine-derived immunity [19–21], and seasonal variation in climatological factors, such as absolute humidity and temperature, that affect viral survival and transmissibility [22–24, 27]. Behavioral changes, including altered contact patterns during holidays [25, 26, 29] or in response to weather conditions [30, 31], further modulate transmission efficiency. Acting together, these mechanisms can generate secondary peaks and abrupt shifts in epidemic growth rates, making accurate forecasting particularly challenging [32].

These intraseason mechanisms are illustrated by the 2017–2018 influenza season, highlighted in red in Fig. A1. The decomposition of hospital admissions for influenza A/H1N1pdm09, influenza A/H3N2, and influenza B reveals how subtype dynamics contribute to the observed variability in the aggregated case counts. In particular, the shaded period corresponding to the first half of January marks a recurrent post-holiday slowdown in epidemic growth that is apparent across multiple seasons [29, 63]. During this interval, a noticeable inflection appears in the trajectory of influenza A/H1N1pdm09, followed by a brief plateau in total incidence driven by the peak of this subtype in the second half of January. However, the overall peak is only reached in early February, coinciding with a rapid increase of influenza A/H3N2 and influenza B. Together, these features illustrate how within-season type replacement and behavioral modulation interact to produce complex, multi-peaked epidemic dynamics, even within a single influenza season.

**Fig. A1.**
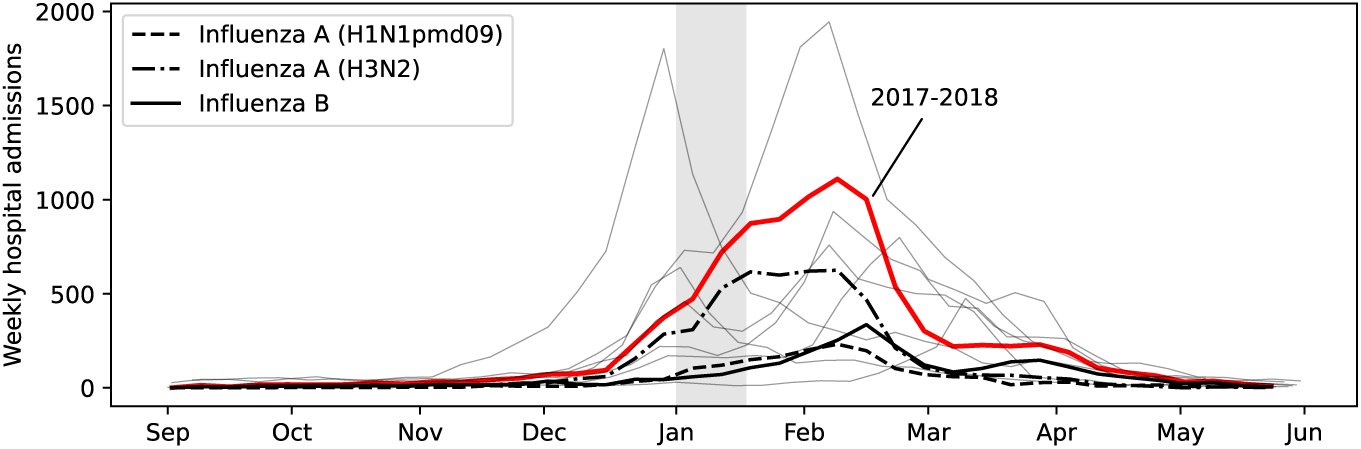
Weekly hospital admissions for influenza in North Carolina from 2014– 2015 until 2024–2025. (excluding SARS-CoV-2 pandemic years 2020–2021, 2021–2022 and 2022–2023). Due to influenza type A/H1N1pdm09 (black, dashed). Due to influenza type A/H3N2 (black, dash-dotted). Due to influenza type B (black, solid). Due to all influenza types, subtypes and lineages combined in season 2017–2018 (red). Due to all influenza types, subtypes and lineages combined in other influenza seasons (thin grey lines). The first half of January (shaded in grey) often coincides with a decrease in admissions. Data sources are listed in Section B.4.

### Appendix B Supplementary methods

#### B.1 Forward simulation model

**Table A1.**
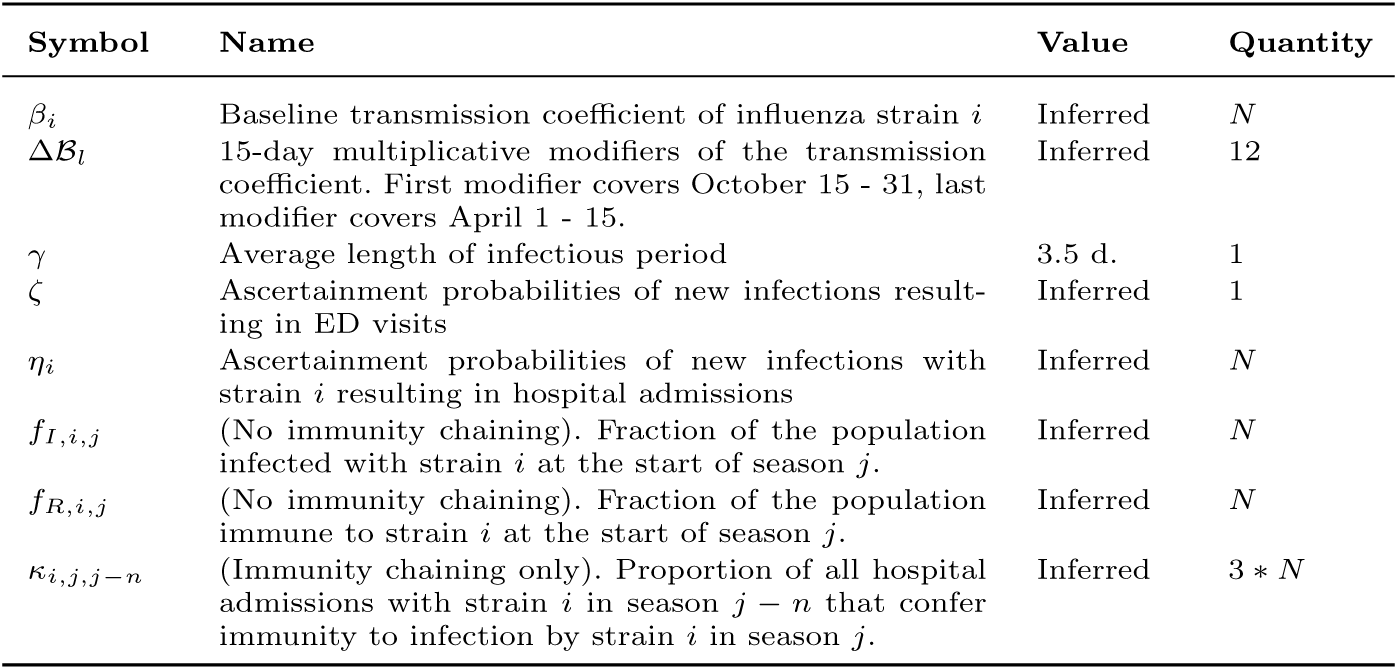
Parameters of the forward simulation model. *N* denotes the number of modeled strains. Without immunity chaining, for *N* = 1, there are 17 unknown parameters whose distributions will be inferred using empirical data (section 4.1.2), while for *N* = 2 and *N* = 3, there are 21 and 25 inferred parameters respectively.

#### B.2 Training and Forecasting

##### Training

The Bayesian hierarchical inference framework’s posterior is,

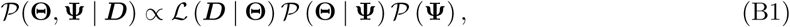

where **Θ** are within-season parameters, **Ψ** are cross-season hyperdistributions which capture cross-seasonal trends and act as priors for the within-season parameters.

*P*(**Θ**, **Ψ** *|* ***D***) is the posterior distribution of the parameters given the training data ***D***, *L*(***D*** *|* **Θ**) is a weighted Poisson likelihood (Eq. (B3), (B4)). *P* (**Θ** *|* **Ψ**) is the probability of observing the within-season parameters given the cross-season hyperparameters and captures the cross-season hierarchy of the model (Eqs. (B5), (B6)). *P* (**Ψ**) are the prior distributions of the cross-season hyper- parameters, which encode pre-existing knowledge about these parameters or enforce shrinkage (Eq. (B7)).

A training dataset ***D*** consists of at least one prior influenza season, each encapsulating multiple time series of data for that season,

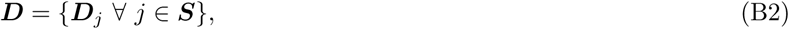

where ***S*** = *{*2014-2015, 2015-2016*, …,* 2024-2025*}* is the set of training seasons and ***D****_j_* = *{*ED_visits_(*t*), H_adm,0_(*t*)*, …,* H_adm*,N*_*_−_*_1_(*t*)*}_j_* is the set of observed time series in season *j*. To simplify the notation in what follows, we represent an individual observed time series of variable *k* in season *j* as *x_j,k_*(*t*) and the corresponding simulation as *y_j,k_*(*t*).

To match the forward simulation to the observed data, we employ a weighted Poisson log-likelihood function for count data, which is normalized such that the weight of an influenza season with an average height of the epidemic peak is one,

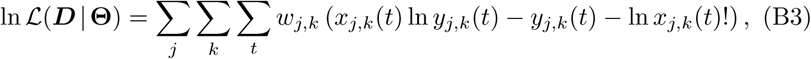

with weights,

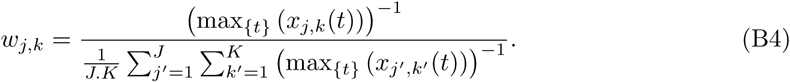

The relationship between the within-season parameters and the cross-season hyperparameters encoded in *P* (**Θ** | **Ψ**) is,

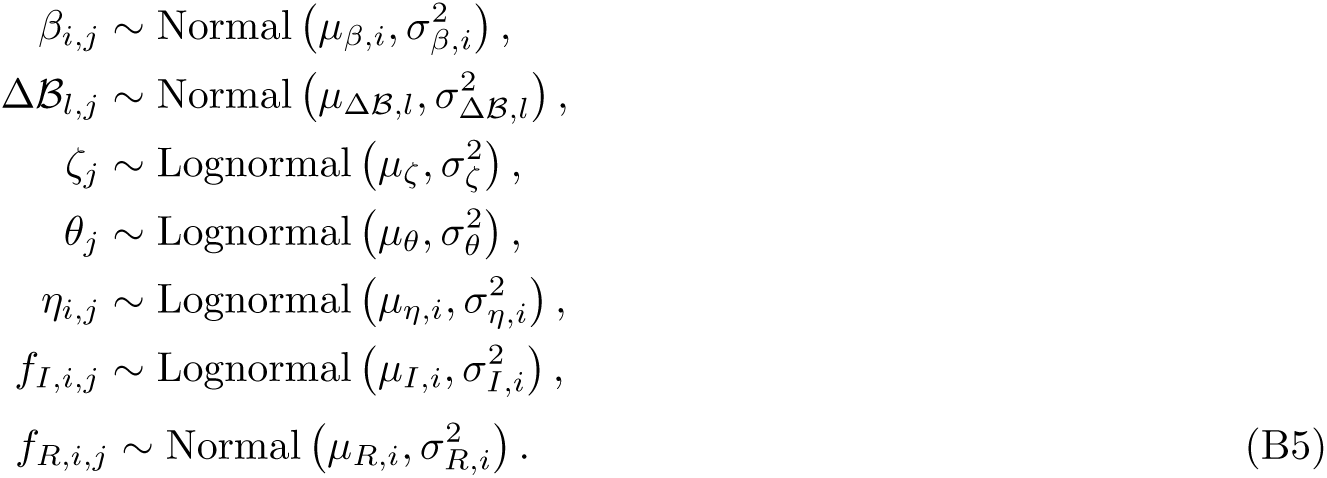

where *i* indexes influenza strains, *j* influenza seasons and *l* the 12 discrete elements of the transmission coefficient modifiers. Lognormal priors were used for the small (*<* 0.05), multiplicative ascertainments (*ζ_j_*, *η_i,j_*) and initial fraction of infected (*f_I,i,j_*), as their uncertainty scales with the parameter. Truncation was applied to the transmission coefficient (*β_i,j_*), initial immunity (*f_R,i,j_*), ascertainments (*ζ_j_*, *η_i,j_*) and initial fraction of infected (*f_I,i,j_*). In practice, none of these parameters approached their physical bounds, so truncation was a conservative formality rather than a modeling necessity. The use of logit- normal distributions were considered, but these proved numerically less stable for small values. If an immunity chaining function is used instead of *f_R,i,j_* (Eqs. (4) and (5)), the parameters *κ_i,j,j−_*_1_, *κ_i,j,j−_*_2_ and *κ_i,j,j−_*_3_ are inferred instead following,

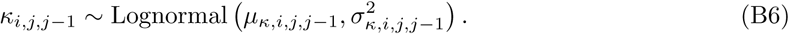

The prior beliefs on the hyperparameters encoded in *P*(**Ψ**) are,

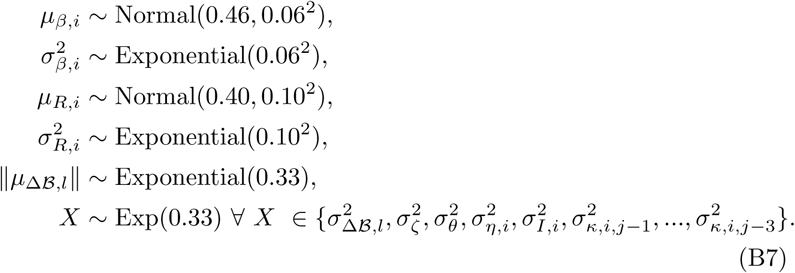

These priors are weakly informative, chosen to stabilize inference by preventing identifiability issues, rather than to strongly constrain parameter estimates. The priors on *µ_β,i_* and 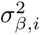 place mass over a broad, historically plausible range of basic reproduction numbers, approximately *R*_0*,i*_ = 1.60 *±* 0.20 [64]. Combined with a prior on population immunity of *f_R,i_* = 0.40 *±* 0.10 [65], the effective reproduction number is placed near unity at the start of a season in the absence of forcing on the transmission coefficient (Δ*β*(0) = 0), providing a centered baseline for epidemic growth. The aim of the priors ∥*µ*_Δ_*_B,l_*∥ and 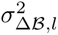 is to shrink the cross-season modifier trajectory toward zero to prevent overfitting. The aim of the priors on the variance of the lognormal distributions is to gently nudge these distributions towards a bell shape rather than being heavy-tailed. This stabilizes inference and prevents extreme draws that could destabilize the model. All remaining priors are uniform distributions.

#### B.3 Accuracy evaluation and baseline

##### Accuracy metrics

We evaluate the accuracy of our model using the geometric mean relative Weighted Interval Score (WIS) [41, 42, 45, 58]. This is computed by dividing the geometric mean of the absolute WIS scores across our model’s forecasts and subsequently normalizing them by the geometric mean of the absolute WIS scores of a baseline model.

Let *i* = *{*0, 1, 2*, . . ., N −* 1*}* index the weeks for which a forecast is made, and *j* = *{*0, 1, 2, 3*}* denote the four-week forecast horizon. Then, for a model *M* and a baseline *B*, the geometric mean relative WIS over the season is defined as:

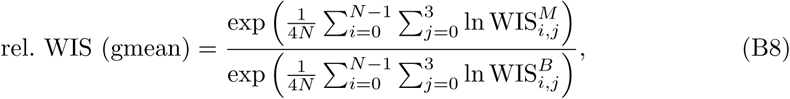

where 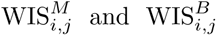 denote the WIS for the model and baseline, respectively, at reference date *i* and forecast horizon *j*.

Alternatively, the arithmetic mean relative WIS score can be computed as follows,

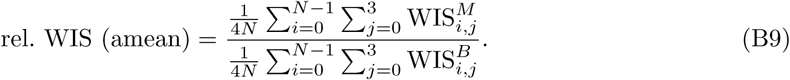

The geometric mean relative WIS weights forecasts more equally and reduces the influence of extreme values. In the context of influenza forecasting, WIS magnitudes increase with forecast horizon [45], and case counts vary substantially across seasons (Fig. A1), and U.S. states and territories (in the context of the CDC FluSight challenge) [46, 47].

Because the geometric mean is not a linear operator, the resulting relative WIS is not a strictly proper scoring metric and may, in principle, incentivize miscalibrated forecasts. CDC FluSight evaluations have historically relied on the arithmetic mean to preserve properness, although organizers have indicated a transition towards the geometric mean (or arithmetic mean of log absolute WIS) for the 2025–2026 season.

In addition to more equally weighing accuracy across U.S. states and territories (in CDC FluSight) and across seasons (in the North Carolina analysis), the geometric mean reduces the relative contribution of longer forecast horizons and downweights epidemic peaks.

Given an observation *y* and a forecast distribution *F*, the WIS is computed as

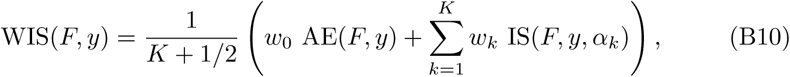

where AE(*F, y*) = *|y − m|* is the absolute error, and IS is the interval score for a single quantile [66]:

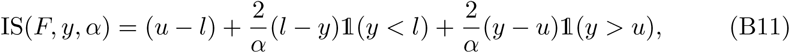

and *l*, *m*, and *u* denote the *α/*2, 0.5 (median), and 1*−α/*2 quantiles of the forecast distribution *F* . Here, 1 is the indicator function. We set *w_k_* = *α_k_/*2 and *w*_0_ = 1*/*2, with *K* = 11 interval scores, for *α* = *{*0.02, 0.05, 0.10, 0.20*, . . .,* 0.90*}* [41, 42, 58]. The WIS rewards narrow prediction intervals and penalizes forecasts that miss the observed value; it is a distributional generalization of the absolute error, and smaller WIS corresponds to higher forecast accuracy.

##### Baseline model

As baseline models, we use two variations of a Geometric Random Walk (GRW), which serve as simple predictors of hospital admissions,

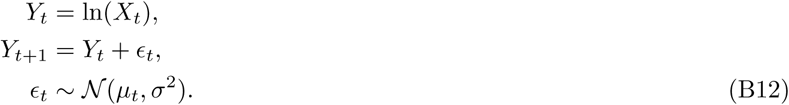

The GRW is implemented in two forms:

1. Stationary. For *µ_t_* = 0, the median of the forecast distribution remains constant over the forecast horizon. A value of *σ* = 0.375 resulted in the lowest WIS across all historical seasons and thus the most accurate stationary GRW model.
2. Non-stationary. Here, the drift *µ_t_* is allowed to vary week-over-week along the four-week forecast horizon, computed from historical data on a leave- one-out basis (Fig. A2). This variant provides a slightly more challenging benchmark for the model.

**Fig. A2.**
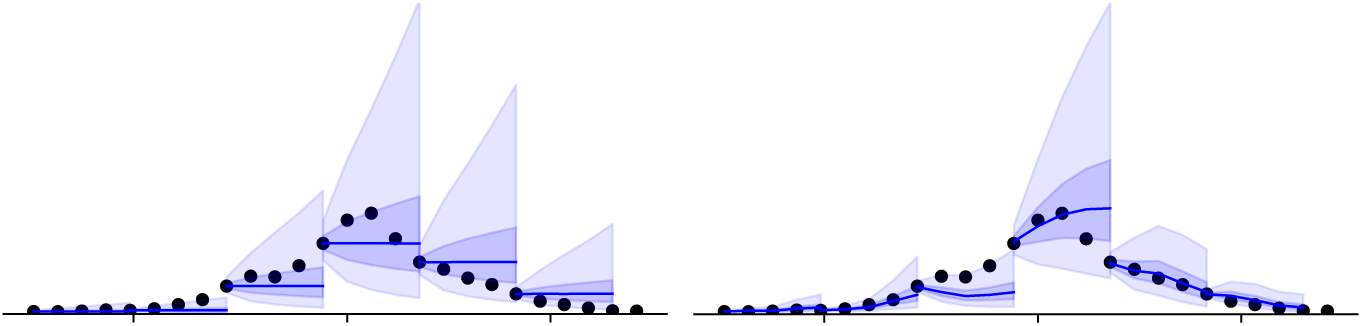
Baseline models used to normalize accuracy. Stationary Geometric Random Walk (left). (right) Non-stationary Geometric Random Walk (right). Weekly influenza hospital admissions due to influenza in North Carolina during the 2024–2025 season (markers).

#### B.4 Available North Carolina data

For eight influenza seasons in North Carolina from 2014-2015 until 2024-2025 (excluding the SARS-CoV-2 pandemic years 2020-2021, 2021-2022, and 2022- 2023), we were able to use four publicly available time series, reported at a weekly frequency:

1. Number of new ED visits with an influenza diagnosis (ICD-9/10-CM, [67]) in NC reported to NC DETECT [68, 69]. ICD diagnosis codes for influenza include 487-488 (all inclusive) for ICD-9-CM and J09-J11 (all inclusive) for ICD-10-CM.
2. Number of new hospital admissions through the ED with an influenza diagnosis (ICD-9/10-CM, [67]) in NC reported to NC DETECT [68, 69].
3. Ratio of tests positive for influenza type A versus influenza type B reported by hospitals in the NC Public Health Epidemiologist Network [70].
4. Ratio of tests positive for influenza A/H1N1pdm09 versus A/H3N2 reported to the Centers of Disease Control and Prevention (CDC) by Public Health Laboratories in the South HHS region [71].

To construct four time series matching the observed states from the disease transmission model (Eq. (2)): (i) The number of new ED visits, (ii) the number of new hospital admissions for influenza type A/H1N1pdm09, (iii) the number of new hospital admissions for influenza type A/H3N2 and (iv) the number of new hospital admissions for influenza type B. Distinguishing between influenza type B lineages Victoria and Yamagata was not possible due to the limited number of samples testing positive for influenza B. The hospital admissions data are shown in Figs. A10 and A11.

#### B.5 Influence of model configuration on accuracy

##### B.5.1 Paired Bootstrap of Relative WIS

To provide a nonparametric assessment of forecast performance differences across model configurations, we conducted a paired bootstrap analysis of geometric mean relative WIS. Forecasts were paired by reference date to ensure that differences in forecast skill were evaluated conditional on shared epidemic context and forecast difficulty. Reference dates were then resampled with replacement, and for each bootstrap sample, the geometric mean relative WIS across all forecast horizons and observations was computed as outlined in Section B.3.

For each bootstrap replicate, we computed the mean log-difference between the model configurations. Percent differences in geometric mean relative WIS were then obtained by exponentiating the bootstrap means,

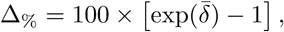

where *δ̄* denotes the mean log-difference between two model configurations. Two-sided 95% confidence intervals were derived from the 2.5th and 97.5th percentiles of the bootstrap distribution. Empirical *p*-values were computed as the proportion of bootstrap replicates with a sign opposite to the observed effect.

##### B.5.2 Linear Mixed-Effects Model of Relative WIS

We fit a linear mixed-effects regression model to the natural logarithm of the absolute WIS obtained from the leave-one-out cross-validation experiment (Sections 4.2.1, B.3). This analysis was restricted to forecasts generated using historically informed forecasts (trained), as the bootstrap analysis demonstrated a large and consistent reduction in relative WIS for these models (Table 1).

Each observation corresponds to a four-week-ahead probabilistic forecast started at a given reference date. Modeling the absolute WIS on the log scale ensures consistency with homoscedasticity and normality assumptions in the linear mixed-effects model’s residual. This choice is also consistent with our use of the geometric mean relative WIS in the paired bootstrap analysis.

We fit the following additive structure,

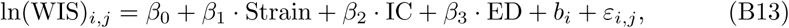

where WIS*_i,j_* denotes the WIS at reference date *i* and forecast horizon *j*. *Strain* is a categorical indicator for the strain representation (1, 2 or 3 strains), *IC* indicates whether immunity chaining was used, and *ED* indicates wether ED visit data were included as an auxiliary signal. A random intercept *b_i_ ∼ N* (0*, σ*^2^) was included for the forecast reference date to account for shared forecast difficulty and outcome variability across models evaluated on the same date.

Model parameters were estimated by restricted maximum likelihood in R using the lme library. We verified that residuals and random effects were approximately normally distributed (Fig. A5). The estimated variance of the reference-date random intercept (1.08) was substantially larger than the residual variance (0.40), indicating that variation in forecast accuracy is dominated by differences across reference dates. This supports the inclusion of reference-date–specific random intercepts to capture substantial heterogeneity in forecast difficulty across epidemic contexts. The marginal *R*^2^ was 0.001 and the conditional *R*^2^ was 0.728, further indicating that while the fixed effects explain only a small fraction of the variability in forecast accuracy, a large proportion is accounted for by reference-date heterogeneity.

#### B.6 Influence of training history on accuracy

##### B.6.1 Non-Linear Mixed-Effects Learning Model

We modeled the dependence of forecast accuracy on the length of the training dataset using the following nonlinear equation on the natural logarithm of absolute WIS,

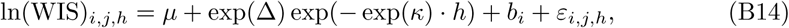

where WIS*_i,j,h_*denotes the WIS at reference date *i* and forecast horizon *j*, made using a model trained on *h* historical seasons.

The nonlinear formulation decomposes learning behavior into three interpretable components. The parameter *µ* represents the asymptotic accuracy achieved in the limit of infinite training data. The parameter Δ captures the accuracy penalty at zero training relative to the asymptotic level, it therein represents the maximum achievable accuracy gain by training. The parameter *κ* governs the rate at which forecast accuracy approaches its asymptotic level as the number of training seasons increases, with larger values indicating faster learning. We model Δ and *κ* on the log scale to ensure these parameters remain strictly positive.

A random intercept *b_i_ ∼ N* (0*, σ*^2^) was included to account for shared epidemic context and forecast difficulty across models evaluated on the same reference date, ensuring that inference is driven by within–reference-date contrasts.

We allowed each component of the learning curve to depend on model configuration through the following additive fixed-effects structure,

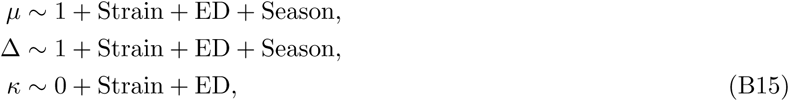

where *Strain* is a categorical indicator for the strain representation (1, 2 or 3 strains), *ED* indicates wether auxiliary ED visits data were included, and *Season* indicates whether the focal season was 2019–2020, 2023–2024 or 2024– 2025.

We fitted the model using the NLME library in R. We confirmed that both residuals and random effects were approximately normally distributed, and that residual variance did not depend on the training horizon (Fig. A6). The estimated variances of the residual error and reference-date random effects were of comparable magnitude, supporting the inclusion of reference-date–specific random intercepts to capture heterogeneity in forecast difficulty across epidemic contexts.

#### B.7 Participation in the CDC FluSight Challenge

##### B.7.1 Underreporting model

###### Overview

The CDC FluSight modeling challenge [46, 47] relies on the National Healthcare Safety Network (NHSN) Hospital Respiratory Dataset (HRD) [62] to provide timely counts of lab-confirmed influenza hospital admissions. Preliminary versions of epidemiological week *X*’s data are first released on Wednesday and later consolidated on Friday of week *X* + 1. FluSight forecasts are generated using the Wednesday preliminary data release, which becomes available at noon with submissions due by 11 pm EST/EDT the same day. A key challenge for accurate forecasting is that these data are systematically incomplete at the time of release, with hospitalization counts revised upward over subsequent weeks as delayed reports are incorporated. We use a generalized Dirichlet–multinomial reporting model that represents reporting as a sequential beta-binomial survival process [59, 61], in which admissions are reported immediately, after one week, or after two weeks with state-specific probabilities. Reporting completeness is estimated from historical revision patterns and uses conjugate Bayesian updating to analytically compute posterior means and avoid sampling. The approach operates at the state level and uses a rolling window of recent data releases [60, 61], allowing it to adapt to evolving reporting behavior while retaining closed-form analytical inference.

##### Data structure

Let *X* denote the epidemiological week in which our forecast is due and consider three consecutive preliminary data releases. For a given US state, define *y_{X−_*_1*,X*_*_}_* as the hospital admissions in week *X−*1 reported in the preliminary dataset of week *X*, *y_{X−_*_1*,X*+1_*_}_* as the hospital admissions in week *X −* 1 reported in week *X* + 1, and *y_{X−_*_1*,X*+2_*_}_* as the hospital admissions in week *X −* 1 reported in week *X* + 2. We assume that *y_{X−_*_1*,X*+2_*_}_* is effectively final. Reporting increments, representing admissions first reported with delays of 0, 1, and 2 weeks, are defined as

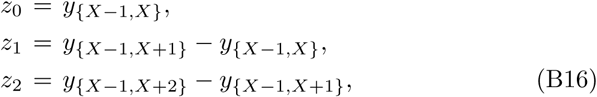

To stabilize inference, reporting increments are aggregated over a rolling window of four weeks for each state. Under the assumption that reporting- delay mechanisms are approximately constant within this window, aggregating weekly increments is equivalent to pooling independent realizations of the same reporting process, thereby increasing effective sample size and stabilizing posterior estimates. The rolling window therefore increases robustness while remaining adaptive to changes in reporting behavior. It represents a deliberate compromise between modeling flexibility and analytical tractability: while calendar-specific reporting effects could be modeled using fully time-varying delay processes, doing so would generally preclude closed-form conjugate inference and require simulation-based methods which represent an operational breakpoint in a real-time forecasting setting.

#### Probabilistic model and posterior inference

Conditional on the finalized admission count *n*, reporting is modeled as a sequential process. Let *v*_0_ denote the probability that an admission is reported immediately, and *v*_1_ the probability that an admission not reported immediately is reported after one additional week. Reporting increments are modeled as,

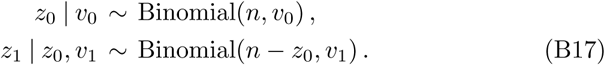

Independent Beta priors are placed on the sequential reporting hazards,

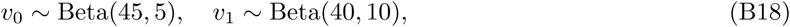

with hyperparameters chosen to encode the prior belief that on average, 90 % of hospital admissions are immediately reported and 98 % are reported after one additional week. By conjugacy [72–74], the posterior distributions are, ’

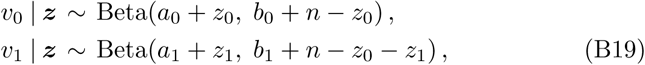

with posterior means available in closed form,

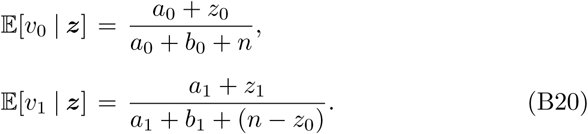

#### Backfilling procedure

Let *y_{X−_*_1*,X*_*_}_* denote the most recent preliminary observation and *y_{X−_*_2*,X*_*_}_* the second-most-recent observation available at release week *X*. These are backfilled to approximate finalized counts via

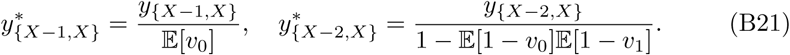

This guarantees monotone completeness,

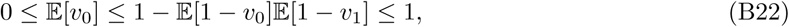

and yields state-specific corrections grounded in recent reporting dynamics.

#### Operational considerations

Of the 53 modeled territories, Arizona, New Jersey, Pennsylvania, and Puerto Rico exhibited net retractions rather than backfill during the 2025–2026 CDC FluSight Challenge. We assumed there was no backfill for these jurisdictions for the sake of operational simplicity. A key strength of the proposed underreporting model is that the posterior means of the reporting hazards remain well-defined and appropriately shrunk toward the prior when there is retraction. From an engineering perspective, the model acts as a noise filter, with the concentrations of the Beta priors governing filtering strength and the rolling window length controlling responsiveness to changes in the reporting regime changes.

The exploitation of Bayesian conjugacy yields a closed-form analytical solution that is computationally inexpensive and avoids sampling-based inference, which represents a potential source of error under operational circumstances.

### Appendix C Supplementary results

#### C.1 Influence of model configuration on accuracy

**Fig. A3.**
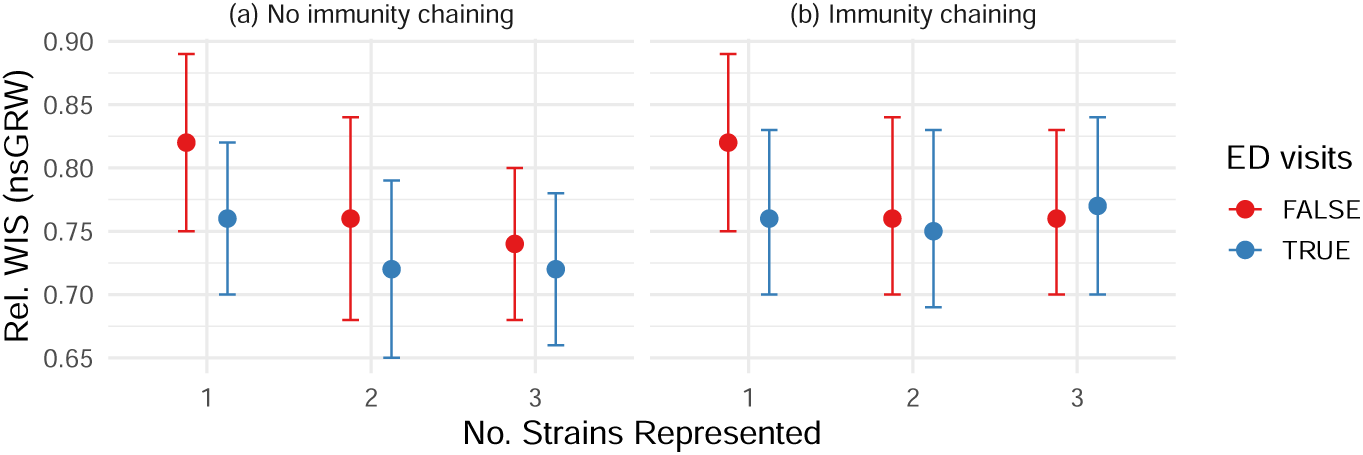
Accuracy of 12 model variants across eight NC influenza seasons. (Section 4.2.1). Lower values correspond to more accurate forecasts. The inclusion of ED visits improved accuracy, and so did the inclusion of strain granularity, albeit with diminishing gains. Accuracy is expressed as the geometric mean Weighted Interval Score relative to a non-stationary Geometric Random Walk baseline model (Section B.3, Fig. 1). The paired bootstrapped median and 95% quantiles are shown (Supplementary Section B.5).

**Fig. A4.**
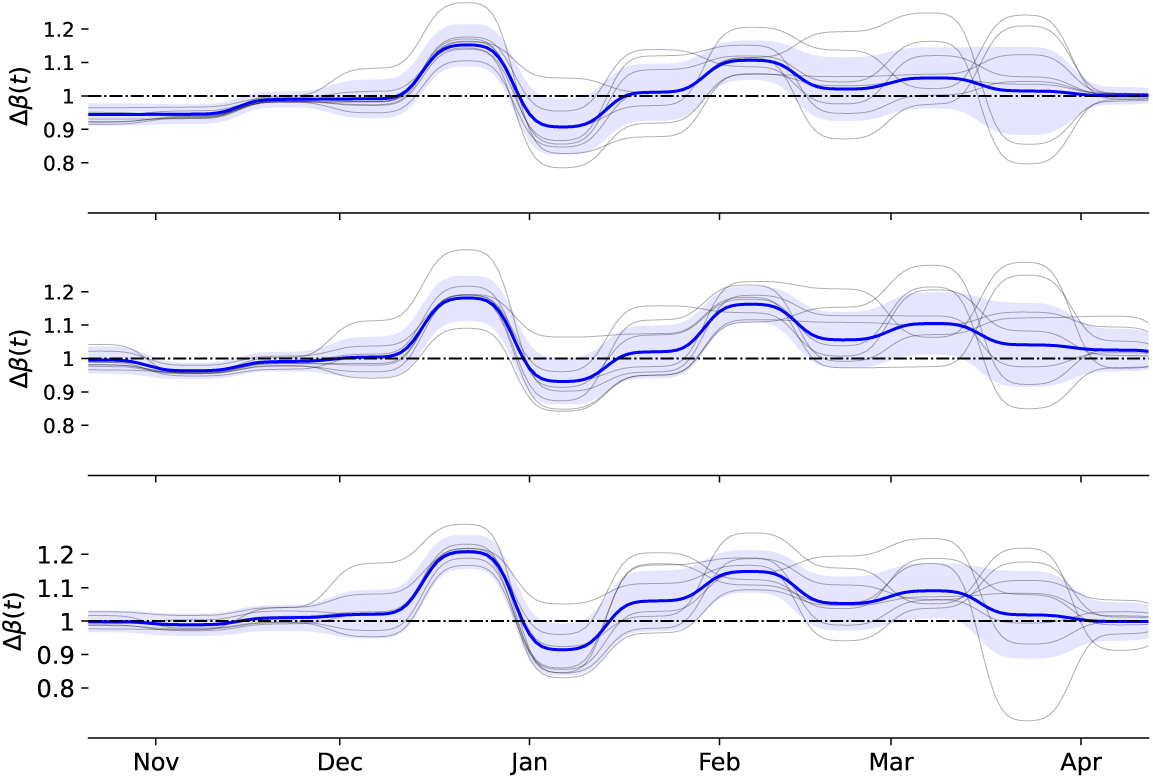
Posterior distribution of the transmission rate modifiers Δ*β*(*t*). Trend across the training seasons with one-sigma confidence interval (blue). Individual training seasons (grey). For the one-strain (aggregated) model (top panel). For the two-strain model distinguishing influenza A and B (middle panel). For the three-strain model distinguishing H1N1pdm09, H3N2, and influenza B (bottom panel). Modifiers show pronounced seasonal structure with a strong post-holiday reduction in transmission. Without immunity chaining and using the ED visits data stream.

**Table A2.** Regression coefficients of the linear mixed-effects model of forecast accuracy (Eq. (B13)). The analysis is restricted to models with informative priors (“Trained”). The reference category for strain representation is the single-strain model.

| Coefficient | Est. | SE | *p*-value |
| --- | --- | --- | --- |
| Intercept | 3.665 | 0.082 | < 0.001 |
| Immunity chaining | +0.024 | 0.014 | 0.088 |
| ED visits | −0.040 | 0.014 | 0.005 |
| 2 Strains | −0.053 | 0.017 | 0.002 |
| 3 Strains | −0.053 | 0.017 | 0.002 |

**Fig. A5.**
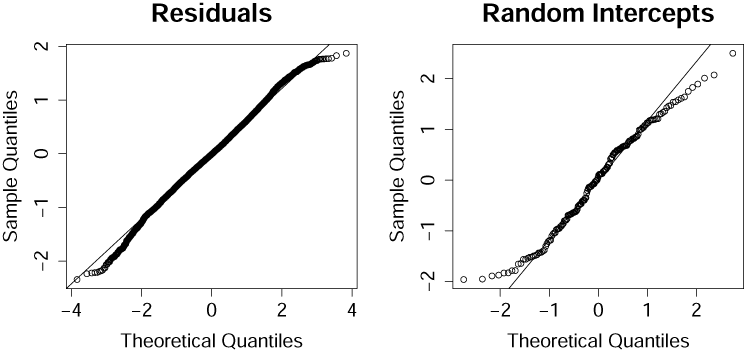
Quantile-quantile plots of the residuals and random effects of the linear mixed-efects model of forecast accuracy (Eq. (B13)).

**Table A3.** Accuracy of all model variants. Geometric mean WIS relative to the stationary and non-stationary GRW baseline models, for the 12 model variants and under both informative and uninformative priors (“Trained”). Obtained from the leave-one-out cross-validation experiment across eight NC influenza seasons (Section 4.2.1). Values represent the paired bootstrapped median and 95% uncertainty intervals of the geometric mean relative WIS (Supplementary Section B.5).

| Trained | Strains | Imm. chain. | ED visits | Rel. WIS (sGRW) | Rel. WIS (nsGRW) |
| --- | --- | --- | --- | --- | --- |
| × | 1 | × | × | 0.74 (0.68–0.81) | 1.00 (0.91–1.10) |
| × | 1 | × | ✓ | 0.67 (0.61–0.74) | 0.91 (0.82–1.01) |
| × | 1 | ✓ | × | 0.77 (0.70–0.85) | 1.04 (0.95–1.15) |
| × | 1 | ✓ | ✓ | 0.68 (0.62–0.75) | 0.92 (0.83–1.02) |
| × | 2 | × | × | 0.72 (0.66–0.79) | 0.98 (0.88–1.08) |
| × | 2 | × | ✓ | 0.71 (0.65–0.79) | 0.96 (0.86–1.08) |
| × | 2 | ✓ | × | 0.73 (0.66–0.80) | 0.98 (0.88–1.09) |
| × | 2 | ✓ | ✓ | 0.70 (0.63–0.77) | 0.94 (0.84–1.04) |
| × | 3 | × | × | 0.76 (0.69–0.83) | 1.03 (0.92–1.14) |
| × | 3 | × | ✓ | 0.73 (0.66–0.80) | 0.99 (0.88–1.10) |
| × | 3 | ✓ | × | 0.74 (0.68–0.82) | 1.01 (0.91–1.12) |
| × | 3 | ✓ | ✓ | 0.77 (0.69–0.85) | 1.01 (0.90–1.14) |
| ✓ | 1 | × | × | 0.61 (0.54–0.67) | 0.82 (0.75–0.89) |
| ✓ | 1 | × | ✓ | 0.56 (0.51–0.62) | 0.76 (0.70–0.82) |
| ✓ | 1 | ✓ | × | 0.61 (0.55–0.67) | 0.82 (0.75–0.89) |
| ✓ | 1 | ✓ | ✓ | 0.56 (0.51–0.62) | 0.76 (0.70–0.83) |
| ✓ | 2 | × | × | 0.56 (0.51–0.63) | 0.76 (0.68–0.84) |
| ✓ | 2 | × | ✓ | 0.53 (0.48–0.59) | 0.72 (0.65–0.79) |
| ✓ | 2 | ✓ | × | 0.56 (0.51–0.63) | 0.76 (0.70–0.84) |
| ✓ | 2 | ✓ | ✓ | 0.56 (0.50–0.62) | 0.75 (0.69–0.83) |
| ✓ | 3 | × | × | 0.55 (0.50–0.60) | 0.74 (0.68–0.80) |
| ✓ | 3 | × | ✓ | 0.53 (0.48–0.58) | 0.72 (0.66–0.78) |
| ✓ | 3 | ✓ | × | 0.56 (0.51–0.63) | 0.76 (0.70–0.83) |
| ✓ | 3 | ✓ | ✓ | 0.57 (0.51–0.63) | 0.77 (0.70–0.84) |

#### C.2 Influence of training history on accuracy

**Fig. A6.**
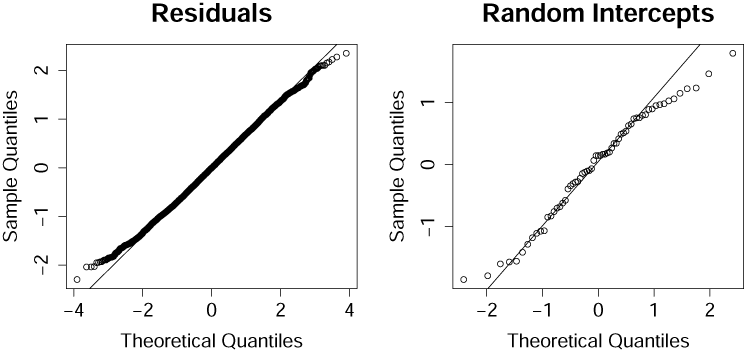
Quantile-quantile plot of the residuals and random effects of the non- linear mixed-efects learning model (Eqs. (B14), (B15)).

**Table A4.**
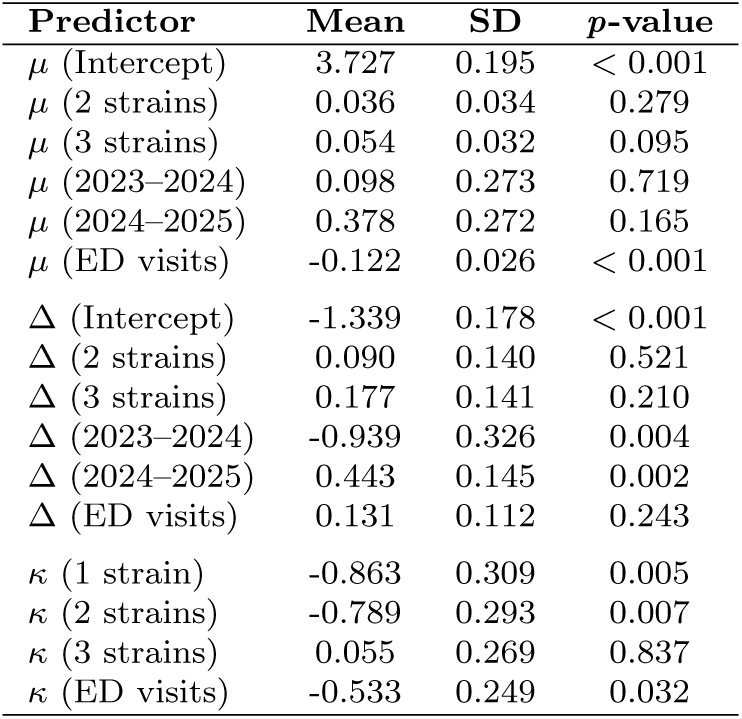
Regression coefficients of the nonlinear mixed-effects learning model (Eqs. (B14), (B15)). Parameters represent asymptotic accuracy (*µ*), accuracy gained by training (exp(Δ)), and training rate (exp(*κ*)). The intercept of *µ* is lower than the mean ln WIS of the stationary GRW (4.714) and non-stationary GRW (4.267) baseline models, indicating our models outperform the baseline models given sufficient training data.

| Predictor | Mean | SD | p-value |
| --- | --- | --- | --- |
| $\mu$ (Intercept) | 3.727 | 0.195 | < 0.001 |
| $\mu$ (2 strains) | 0.036 | 0.034 | 0.279 |
| $\mu$ (3 strains) | 0.054 | 0.032 | 0.095 |
| $\mu$ (2023–2024) | 0.098 | 0.273 | 0.719 |
| $\mu$ (2024–2025) | 0.378 | 0.272 | 0.165 |
| $\mu$ (ED visits) | -0.122 | 0.026 | < 0.001 |
| $\Delta$ (Intercept) | -1.339 | 0.178 | < 0.001 |
| $\Delta$ (2 strains) | 0.090 | 0.140 | 0.521 |
| $\Delta$ (3 strains) | 0.177 | 0.141 | 0.210 |
| $\Delta$ (2023–2024) | -0.939 | 0.326 | 0.004 |
| $\Delta$ (2024–2025) | 0.443 | 0.145 | 0.002 |
| $\Delta$ (ED visits) | 0.131 | 0.112 | 0.243 |
| $\kappa$ (1 strain) | -0.863 | 0.309 | 0.005 |
| $\kappa$ (2 strains) | -0.789 | 0.293 | 0.007 |
| $\kappa$ (3 strains) | 0.055 | 0.269 | 0.837 |
| $\kappa$ (ED visits) | -0.533 | 0.249 | 0.032 |

**Fig. A7.**
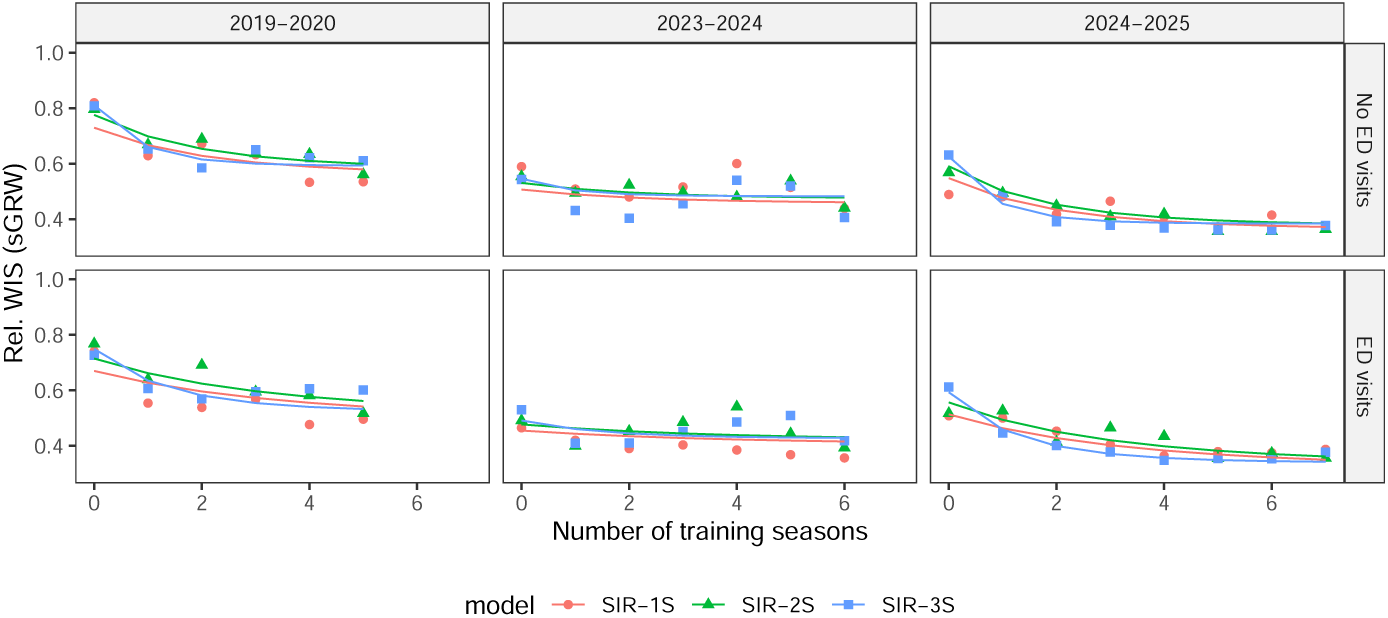
Forecast accuracy in the 2019–2020 (left), 2023–2024 (middle) and 2024–2025 (right) North Carolina influenza seasons as a function of the number of preceding seasons used for training. Lower values correspond to more accurate forecasts. Seasonal context is the dominant determinant of how much training improves performance. Accuracy is expressed as the geometric mean Weighted Interval Score relative to the stationary Geometric Random Walk baseline model (Section B.3, Fig. 3). Markers represent the empirical accuracy, while full lines represent the modeled accuracy (Eqs.(B14), (B15)). Models do not use immunity chaining.

#### C.3 Participation in the CDC FluSight Challenge

##### C.3.1 Drivers of state-level performance

**Fig. A8.**
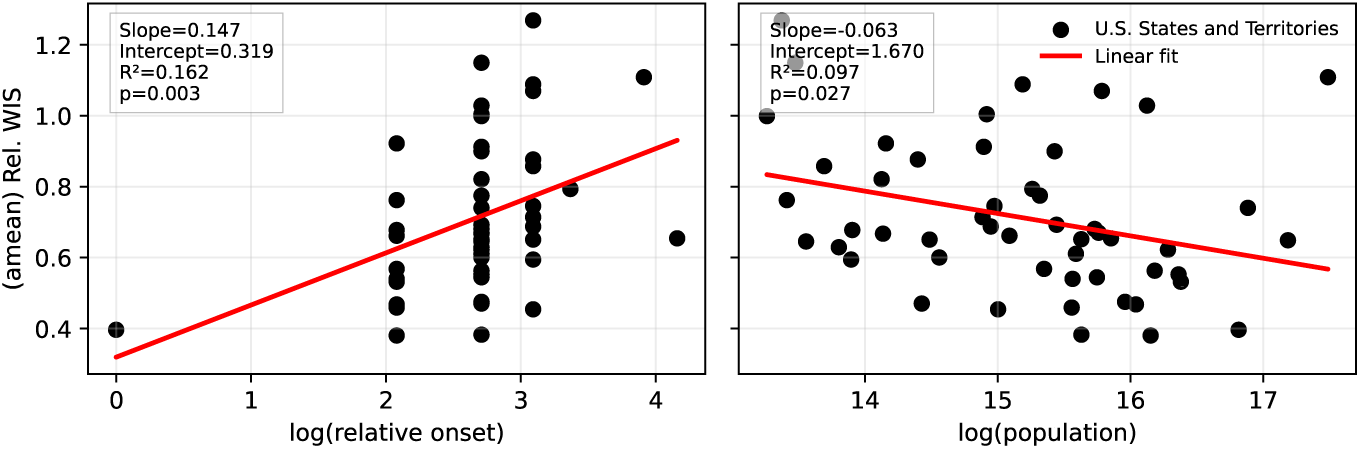
Influence of epidemic onset and population size on state-level 2025–2026 CDC FluSight performance. (left) Arithmetic mean relative WIS versus log-transformed days from earliest epidemic onset, showing a weak-moderate positive association; the epidemic onset date was defined as the first date a U.S. state or territory (excluding Puerto Rico) exceeded 4 hospital admissions per 100,000 inhabitants per week, which occurred on December 6, 2025 in New York. (right) Arithmetic mean relative WIS versus log-transformed population, showing a weak negative association. Each point represents a U.S. state or territory, and regression lines are shown in red.

##### C.3.2 Sensitivity of rankings to forecast omission

To assess the sensitivity of end-of-season model rankings to forecast completeness, we considered the 19 models that submitted forecasts for all 27 forecast weeks of the 2025–2026 CDC FluSight Challenge, which included our model. We then repeated the ranking procedure after omitting one of the 27 forecast weeks from our model’s submissions, resulting in 96.7 % forecast completeness (the other models retaining 100 % completeness). This procedure was repeated separately for each forecast week, and the resulting rankings were calculated using both the arithmetic and geometric mean relative WIS (Fig. A9).

Omitting a single forecast during the period from early December 2025 to mid- January 2026 improved our model’s end-of-season ranking from third to first based on the arithmetic mean relative WIS, and from tenth to ninth based on the geometric mean relative WIS. In contrast, omitting a forecast from mid-March 2026 through the end of the challenge consistently worsened our model’s ranking.

The epidemic peak is both the most challenging period to forecast and the period during which the end-of-season ranking is most sensitive, because WIS is expressed on the same scale as hospital admissions. Omitting a forecast during this period can therefore reduce the contribution of a difficult forecast to the final score, leading to an improvement in ranking. In contrast, late- season forecasts, after the epidemic peak has passed, are substantially more predictable. Omitting a forecast during this period therefore removes a comparatively favorable forecast from the score, resulting in a deterioration of the ranking.

These results demonstrate that end-of-season rankings can be highly sensitive to forecast completeness. Comparisons between models with different submission rates may therefore not provide a fair representation of their relative forecasting skill, as models with incomplete submissions can effectively exclude particularly challenging forecast periods from their final score. More broadly, this creates an opportunity for strategic omission of forecasts to improve a model’s ranking, even when rankings are based on a proper scoring rule.

**Fig. A9.**
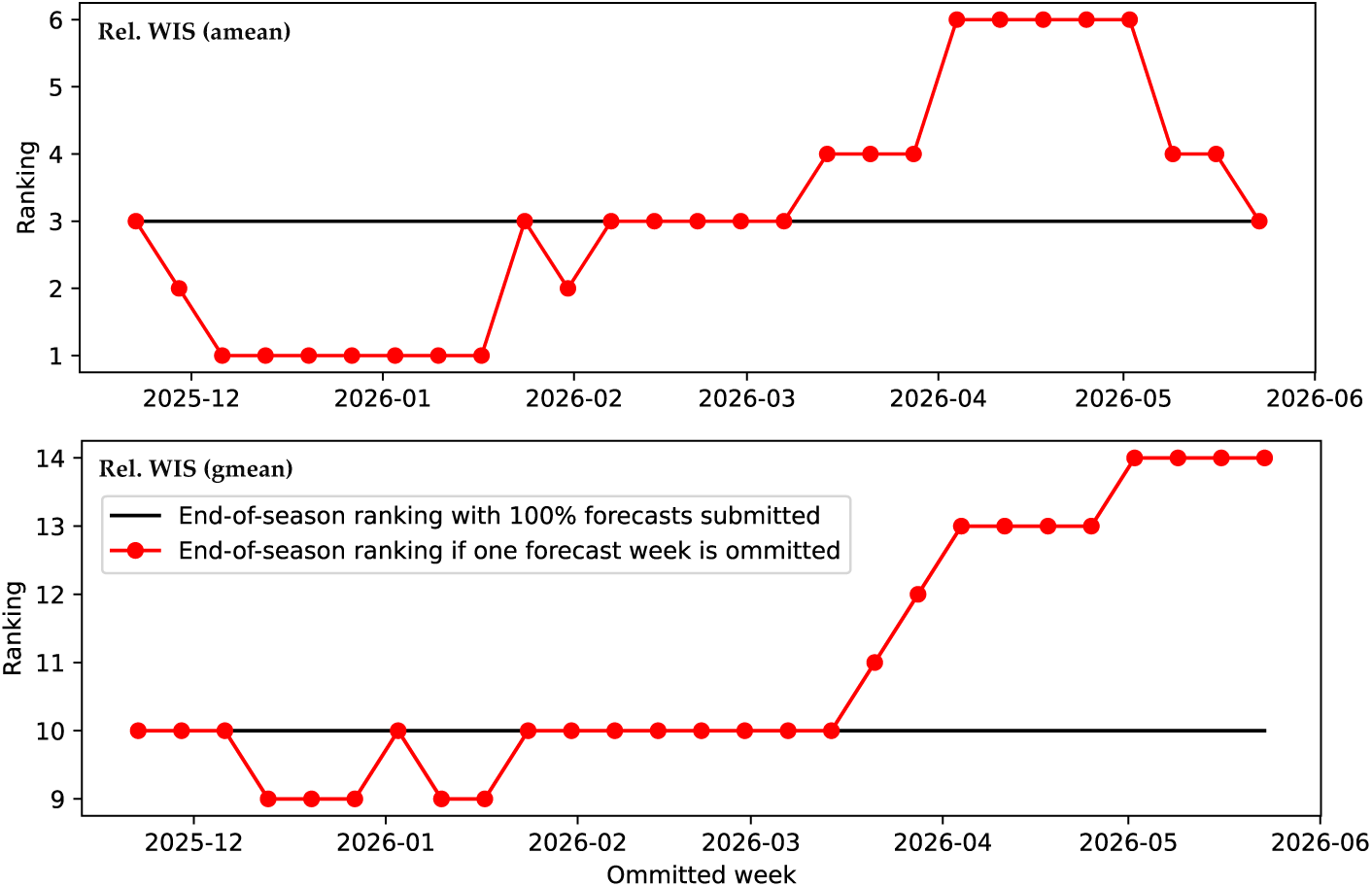
Sensitivity of our model’s ranking to the omission of one forecast week. (top) Sensitivity of the arithmetic mean relative WIS. (bottom) Sensitivity of the geometric mean realtive WIS.

### Appendix D North Carolina Influenza Data

**Fig. A10.**
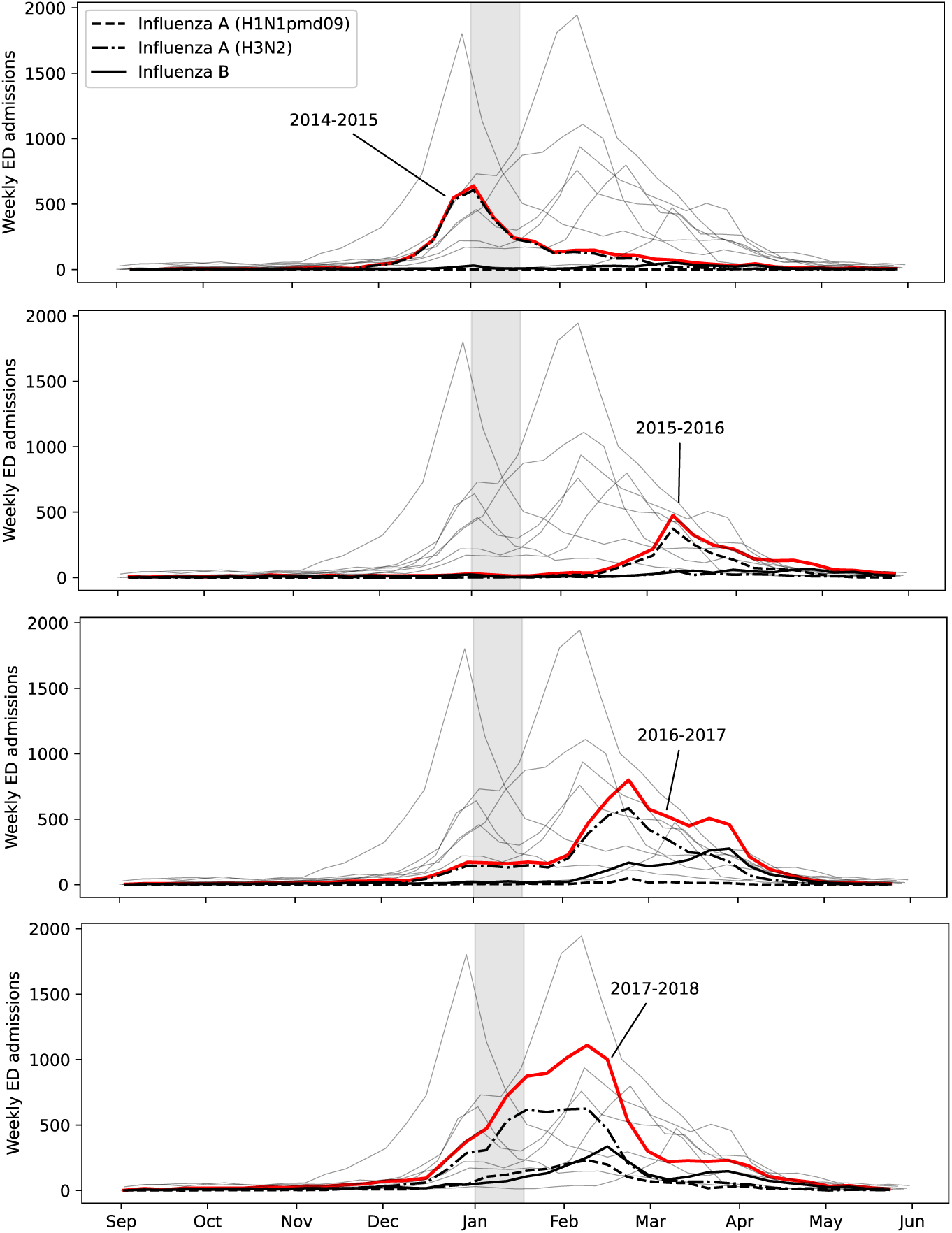
Weekly hospital admissions for influenza in North Carolina. Due to influenza type A/H1N1pdm09 (black, dashed). Due to influenza type A/H3N2 (black, dash-dotted). Due to influenza type B (black, solid). Due to all influenza types, subtypes and lineages combined (red). Due to all influenza types, subtypes and lineages combined in other influenza seasons (thin grey lines). The first half of January (shaded in grey) often coincides with a decrease in admissions. Data sources are listed in Section B.4.

**Fig. A11.**
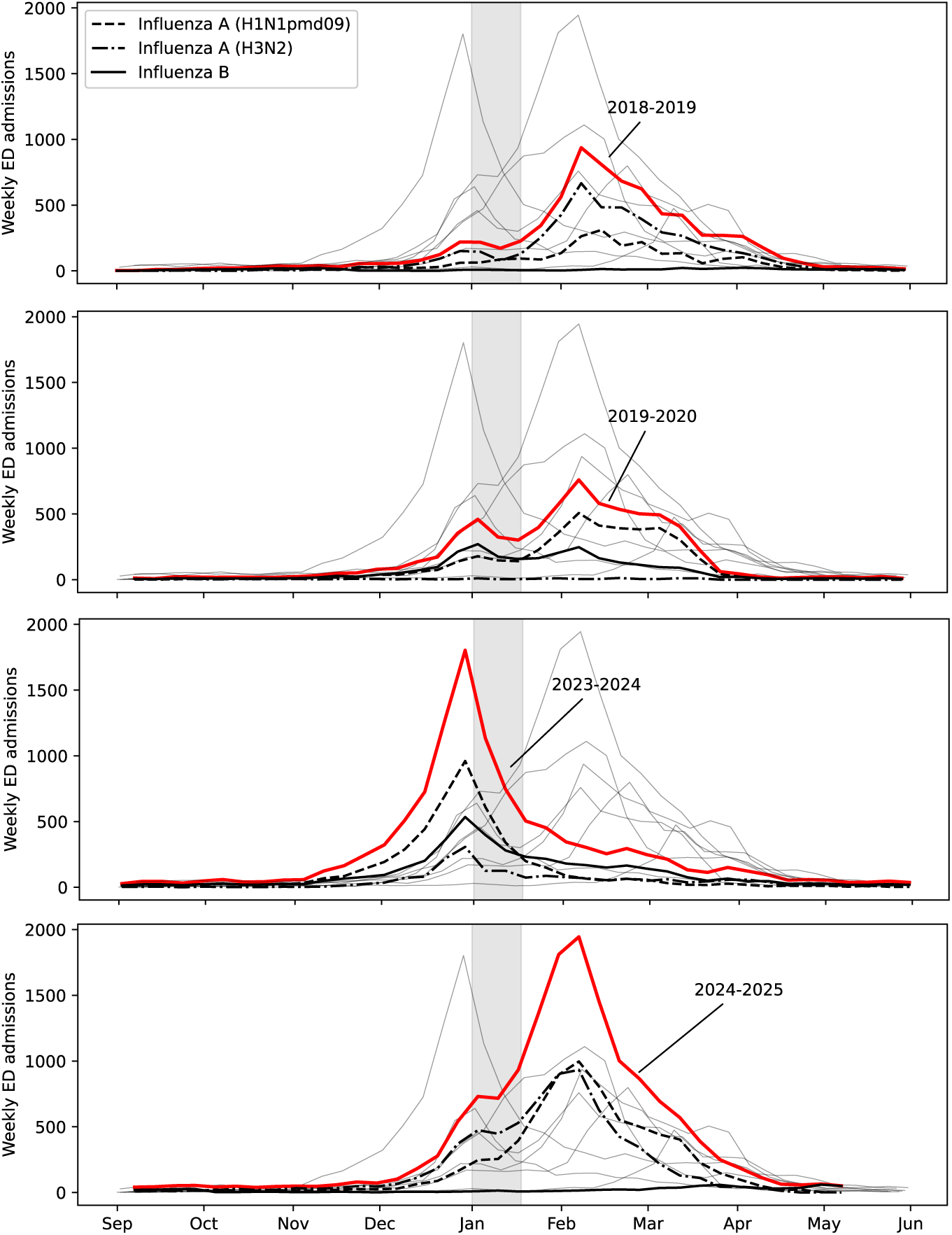
Weekly hospital admissions for influenza in North Carolina. Due to influenza type A/H1N1pdm09 (black, dashed). Due to influenza type A/H3N2 (black, dash-dotted). Due to influenza type B (black, solid). Due to all influenza types, subtypes and lineages combined (red). Due to all influenza types, subtypes and lineages combined in other influenza seasons (thin grey lines). The first half of January (shaded in grey) often coincides with a decrease in admissions. Data sources are listed in Section B.4.

## References

[1] Krammer, F., Smith, G.J.D., Fouchier, R.A.M., Peiris, M., Kedzierska, K., Doherty, P.C., Palese, P., Shaw, M.L., Treanor, J., Webster, R.G., García-Sastre, A.: Influenza. Nature Reviews Disease Primers 4(1), 3 (2018). 10.1038/s41572-018-0002-y

[2] Iuliano, A.D., Roguski, K.M., Chang, H.H., Muscatello, D.J., Palekar, R., Tempia, S., Cohen, C., Gran, J.M., Schanzer, D., Cowling, B.J., Wu, P., Kyncl, J., Ang, L.W., Park, M., Redlberger-Fritz, M., Yu, H., Espenhain, L., Krishnan, A., Emukule, G., van Asten, L., Pereira da Silva, S., Aungkulanon, S., Buchholz, U., Widdowson, M.-A., Bresee, J.S., Azziz-Baumgartner, E., Cheng, P.-Y., Dawood, F., Foppa, I., Olsen, S., Haber, M., Jeffers, C., MacIntyre, C.R., Newall, A.T., Wood, J.G., Kundi, M., Popow-Kraupp, T., Ahmed, M., Rahman, M., Marinho, F., Sotomayor Proschle, C.V., Vergara Mallegas, N., Luzhao, F., Sa, L., Barbosa-Ramírez, J., Sanchez, D.M., Gomez, L.A., Vargas, X.B., Acosta Herrera, a., Llanés, M.J., Fischer, T.K., Krause, T.G., Mølbak, K., Nielsen, J., Trebbien, R., Bruno, A., Ojeda, J., Ramos, H., an der Heiden, M., del Carmen Castillo Signor, L., Serrano, C.E., Bhardwaj, R., Chadha, M., Narayan, V., Kosen, S., Bromberg, M., Glatman-Freedman, A., Kaufman, Z., Arima, Y., Oishi, K., Chaves, S., Nyawanda, B., Al-Jarallah, R.A., Kuri-Morales, P.A., Matus, C.R., Corona, M.E.J., Burmaa, A., Darmaa, O., Obtel, M., Cherkaoui, I., van den Wijngaard, C.C., van der Hoek, W., Baker, M., Bandaranayake, D., Bissielo, A., Huang, S., Lopez, L., Newbern, C., Flem, E., Grøneng, G.M., Hauge, S., de Cosío, F.G., de Moltó, Y., Castillo, L.M., Cabello, M.A., von Horoch, M., Medina Osis, J., Machado, A., Nunes, B., Rodrigues, A.P., Rodrigues, E., Calomfirescu, C., Lupulescu, E., Popescu, R., Popovici, O., Bogdanovic, D., Kostic, M., Lazarevic, K., Milosevic, Z., Tiodorovic, B., Chen, M., Cutter, J., Lee, V., Lin, R., Ma, S., Cohen, A.L., Treurnicht, F., Kim, W.J., Delgado-Sanz, C., de mateo Ontañón, S., Larrauri, A., León, I.L., Vallejo, F., Born, R., Junker, C., Koch, D., Chuang, J.-H., Huang, W.- T., Kuo, H.-W., Tsai, Y.-C., Bundhamcharoen, K., Chittaganpitch, M., Green, H.K., Pebody, R., Goñi, N., Chiparelli, H., Brammer, L., Mus-taquim, D.: Estimates of global seasonal influenza-associated respiratory mortality: a modelling study. The Lancet 391(10127), 1285–1300 (2018). 10.1016/S0140-6736(17)33293-2

[3] World Health Organization: Fact Sheets: Influenza (seasonal) (2025). https://www.who.int/news-room/fact-sheets/detail/influenza-(seasonal) Accessed 2026-01-30

[4] Koelle, K., Cobey, S., Grenfell, B., Pascual, M.: Epochal evolution shapes the phylodynamics of interpandemic influenza a (h3n2) in humans. Science 314(5807), 1898–1903 (2006). 10.1126/science.1132745

[5] Bedford, T., Suchard, M.A., Lemey, P., Dudas, G., Gregory, V., Hay, A.J., McCauley, J.W., Russell, C.A., Smith, D.J., Rambaut, A.: Integrating influenza antigenic dynamics with molecular evolution. eLife 3, 01914 (2014). 10.7554/eLife.01914

[6] Cobey, S., Hensley, S.E.: Immune history and influenza virus susceptibility. Current Opinion in Virology 22, 105–111 (2017). 10.1016/j.coviro.2016.12.004. Emerging viruses: intraspecies transmission • Viral immunology

[7] Hill, E.M., Petrou, S., de Lusignan, S., Yonova, I., Keeling, M.J.: Seasonal influenza: Modelling approaches to capture immunity propagation. PLOS Computational Biology 15(10), 1–26 (2019). 10.1371/journal.pcbi.1007096

[8] Centers for Disease Control and Prevention: Past Seasons’ Vaccine Effectiveness Estimates (2025). https://www.cdc.gov/flu-vaccines-work/php/effectiveness-studies/past-seasons-estimates.html Accessed 2026-01-30

[9] Kawai, N., Ikematsu, H., Bando, T., Kawashima, T., Matsuura, S., Maeda, T., Lee, W.J., Nagao, S., Yoshimura, M., Mori, K., Tanaka, O., Doniwa, K.-i., Satoh, I., Kashiwagi, S.: Influenza vaccine effectiveness over 17 seasons in a large japanese cohort: Analyses by age, virus type, underlying diseases and seasons before the covid-19 pandemic. Journal of Infection and Public Health 18(11), 102934 (2025). 10.1016/j.jiph.2025.102934

[10] Ranjeva, S., Subramanian, R., Fang, V.J., Leung, G.M., Ip, D.K.M., Perera, R.A.P.M., Peiris, J.S.M., Cowling, B.J., Cobey, S.: Age-specific differences in the dynamics of protective immunity to influenza. Nature Communications 10(1), 1660 (2019). 10.1038/s41467-019-09652-6

[11] Arevalo, P., McLean, H.Q., Belongia, E.A., Cobey, S.: Earliest infections predict the age distribution of seasonal influenza A cases. eLife 9, 50060 (2020). 10.7554/eLife.50060

[12] Vieira, M.C., Donato, C.M., Arevalo, P., Rimmelzwaan, G.F., Wood, T., Lopez, L., Huang, Q.S., Dhanasekaran, V., Koelle, K., Cobey, S.: Lineage-specific protection and immune imprinting shape the age distributions of influenza B cases. Nature Communications 12(1), 4313 (2021). 10.1038/s41467-021-24566-y

[13] Leung, N.H.L., Xu, C., Ip, D.K.M., Cowling, B.J.: Review article: The fraction of influenza virus infections that are asymptomatic: A systematic review and meta-analysis. Epidemiology 26(6), 862–872 (2015)

[14] Caini, S., Kroneman, M., Wiegers, T., El Guerche-Séblain, C., Paget, J.: Clinical characteristics and severity of influenza infections by virus type, subtype, and lineage: A systematic literature review. Influenza Other Respir Viruses 12(6), 780–792 (2018)

[15] Zhang, C., Huang, X., Fang, V.J., Chan, K.-H., Leung, G.M., Ip, D.K.M., Peiris, J.S.M., Cowling, B.J., Cauchemez, S., Tsang, T.K.: Frequent presymptomatic household transmission of influenza a but not influenza b virus. Nature Health (2026). 10.1038/ s44360-025-00049-y

[16] Chattopadhyay, I., Kiciman, E., Elliott, J.W., Shaman, J.L., Rzhetsky, A.: Conjunction of factors triggering waves of seasonal influenza. eLife 7, 30756 (2018). 10.7554/eLife.30756

[17] Truscott, J., Fraser, C., Cauchemez, S., Meeyai, A., Hinsley, W., Donnelly, C.A., Ghani, A., Ferguson, N.: Essential epidemiological mechanisms underpinning the transmission dynamics of seasonal influenza. Journal of The Royal Society Interface 9(67), 304–312 (2012). 10.1098/rsif.2011.0309

[18] Nickbakhsh, S., Mair, C., Matthews, L., Reeve, R., Johnson, P.C.D., Thorburn, F., von Wissmann, B., Reynolds, A., McMenamin, J., Gunson, R.N., Murcia, P.R.: Virus–virus interactions impact the population dynamics of influenza and the common cold. Proceedings of the National Academy of Sciences 116(52), 27142–27150 (2019). 10.1073/pnas.1911083116

[19] Ferdinands, J.M., Fry, A.M., Reynolds, S., Petrie, J.G., Flannery, B., Jackson, M.L., Belongia, E.A.: Intraseason waning of influenza vaccine protection: evidence from the us influenza vaccine effectiveness network, 2011–2012 through 2014–2015. Clinical Infectious Diseases **64**(5), 544–550 (2017)

[20] Young, B., Zhao, X., Cook, A.R., Parry, C.M., Wilder-Smith, A., I-Cheng, M.C.: Do antibody responses to the influenza vaccine persist year-round in the elderly? a systematic review and meta-analysis. Vaccine 35(2), 212–221 (2017)

[21] Ray, G.T., Lewis, N., Klein, N.P., Daley, M.F., Wang, S.V., Kulldorff, M., Fireman, B.: Intraseason waning of influenza vaccine effectiveness. Clinical Infectious Diseases 68(10), 1623–1630 (2019)

[22] Shaman, J., Pitzer, V.E., Viboud, C., Grenfell, B.T., Lipsitch, M.: Absolute humidity and the seasonal onset of influenza in the continental united states. PLOS Biology 8(2), 1–13 (2010). 10.1371/journal.pbio.1000316

[23] Tamerius, J.D., Shaman, J., Alonso, W.J., Bloom-Feshbach, K., Uejio, C.K., Comrie, A., Viboud, C.: Environmental predictors of seasonal influenza epidemics across temperate and tropical climates. PLOS Pathogens 9(3), 1–12 (2013). 10.1371/journal.ppat.1003194

[24] Axelsen, J.B., Yaari, R., Grenfell, B.T., Stone, L.: Multiannual forecasting of seasonal influenza dynamics reveals climatic and evolutionary drivers. Proceedings of the National Academy of Sciences 111(26), 9538–9542 (2014). 10.1073/pnas.1321656111

[25] Eames, K.T.D., Tilston, N.L., Edmunds, W.J.: The impact of school holidays on the social mixing patterns of school children. Epidemics 3(2), 103–108 (2011). 10.1016/j.epidem.2011.03.003

[26] Béraud, G., Kazmercziak, S., Beutels, P., Levy-Bruhl, D., Lenne, X., Mielcarek, N., Yazdanpanah, Y., Boëlle, P.-Y., Hens, N., Dervaux, B.: The french connection: The first large population-based contact survey in france relevant for the spread of infectious diseases. PLOS ONE 10(7), 1–22 (2015). 10.1371/journal.pone.0133203

[27] Deyle, E.R., Maher, M.C., Hernandez, R.D., Basu, S., Sugihara, G.: Global environmental drivers of influenza. Proceedings of the National Academy of Sciences 113(46), 13081–13086 (2016). 10.1073/pnas.1607747113

[28] Charu, V., Zeger, S., Gog, J., Bjørnstad, O.N., Kissler, S., Simonsen, L., Grenfell, B.T., Viboud, C.: Human mobility and the spatial transmission of influenza in the united states. PLOS Computational Biology 13(2), 1–23 (2017). 10.1371/journal.pcbi.1005382

[29] Ewing, A., Lee, E.C., Viboud, C., Bansal, S.: Contact, travel, and transmission: The impact of winter holidays on influenza dynamics in the united states. J Infect Dis 215(5), 732–739 (2017)

[30] Willem, L., Van Kerckhove, K., Chao, D.L., Hens, N., Beutels, P.: A nice day for an infection? weather conditions and social contact patterns relevant to influenza transmission. PLOS ONE 7(11), 1–7 (2012). 10.1371/journal.pone.0048695

[31] Chan, T.-C., Fu, Y.-C., Hwang, J.-S.: Changing social contact patterns under tropical weather conditions relevant for the spread of infectious diseases. Epidemiol Infect 143(2), 440–451 (2014)

[32] Moran, K.R., Fairchild, G., Generous, N., Hickmann, K., Osthus, D., Priedhorsky, R., Hyman, J., Del Valle, S.Y.: Epidemic forecasting is messier than weather forecasting: The role of human behavior and internet data streams in epidemic forecast. The Journal of Infectious Diseases 214(4) (2016). 10.1093/infdis/jiw375

[33] Chan, L.Y.H., Morris, S., Hassell, N., Marcenac, P., Couture, A., Colon, A., Kniss, K., Budd, A., Biggerstaff, M., Borchering, R.: Understanding spatiotemporal clustering of seasonal influenza in the united states. BMC Infectious Diseases 26(1), 746 (2026). 10.1186/s12879-026-13000-7

[34] Yang, W., Karspeck, A., Shaman, J.: Comparison of filtering methods for the modeling and retrospective forecasting of influenza epidemics. PLOS Computational Biology 10(4), 1–15 (2014). 10.1371/journal.pcbi.1003583.

[35] Andronico, A., Paireau, J., Cauchemez, S.: Integrating information from historical data into mechanistic models for influenza forecasting. PLOS Computational Biology 20(10), 1–17 (2024). 10.1371/journal.pcbi.1012523

[36] Quaghebeur, W., Nopens, I., De Baets, B.: Incorporating unmodeled dynamics into first-principles models through machine learning. IEEE Access 9, 22014–22022 (2021). 10.1109/ACCESS.2021.3055353

[37] Ye, Y., Pandey, A., Bawden, C., Sumsuzzman, D.M., Rajput, R., Shoukat, A., Singer, B.H., Moghadas, S.M., Galvani, A.P.: Integrating artificial intelligence with mechanistic epidemiological modeling: a scoping review of opportunities and challenges. Nature Communications 16(1), 581 (2025). 10.1038/s41467-024-55461-x

[38] Osthus, D., Gattiker, J., Priedhorsky, R., Valle, S.Y.D.: Dynamic Bayesian Influenza Forecasting in the United States with Hierarchical Discrepancy (with Discussion). Bayesian Analysis 14(1), 261–312 (2019). 10.1214/18-BA1117

[39] Flaxman, S., Mishra, S., Gandy, A., Unwin, H.J.T., Mellan, T.A., Coupland, H., Whittaker, C., Zhu, H., Berah, T., Eaton, J.W., Monod, M., Perez-Guzman, P.N., Schmit, N., Cilloni, L., Ainslie, K.E.C., Baguelin, M., Boonyasiri, A., Boyd, O., Cattarino, L., Cooper, L.V., Cucunubá, Z., Cuomo-Dannenburg, G., Dighe, A., Djaafara, B., Dorigatti, I., van Elsland, S.L., FitzJohn, R.G., Gaythorpe, K.A.M., Geidelberg, L., Grassly, N.C., Green, W.D., Hallett, T., Hamlet, A., Hinsley, W., Jeffrey, B., Knock, E., Laydon, D.J., Nedjati-Gilani, G., Nouvellet, P., Parag, K.V., Siveroni, I., Thompson, H.A., Verity, R., Volz, E., Walters, C.E., Wang, H., Wang, Y., Watson, O.J., Winskill, P., Xi, X., Walker, P.G.T., Ghani, A.C., Donnelly, C.A., Riley, S., Vollmer, M.A.C., Ferguson, N.M., Okell, L.C., Bhatt, S., Team, I.C.C.-.R.: Estimating the effects of non-pharmaceutical interventions on covid-19 in europe. Nature 584(7820), 257–261 (2020). 10.1038/s41586-020-2405-7

[40] Reich, N.G., Brooks, L.C., Fox, S.J., Kandula, S., McGowan, C.J., Moore, E., Osthus, D., Ray, E.L., Tushar, A., Yamana, T.K., Biggerstaff, M., Johansson, M.A., Rosenfeld, R., Shaman, J.: A collaborative multiyear, multimodel assessment of seasonal influenza forecasting in the united states. Proceedings of the National Academy of Sciences 116(8), 3146–3154 (2019). 10.1073/pnas.1812594116

[41] Cramer, E.Y., Ray, E.L., Lopez, V.K., Bracher, J., Brennen, A., Rivadeneira, A.J.C., Gerding, A., Gneiting, T., House, K.H., Huang, Y., Jayawardena, D., Kanji, A.H., Khandelwal, A., Le, K., Mühlemann, A., Niemi, J., Shah, A., Stark, A., Wang, Y., Wattanachit, N., Zorn, M.W., Gu, Y., Jain, S., Bannur, N., Deva, A., Kulkarni, M., Merugu, S., Raval, A., Shingi, S., Tiwari, A., White, J., Abernethy, N.F., Woody, S., Dahan, M., Fox, S., Gaither, K., Lachmann, M., Meyers, L.A., Scott, J.G., Tec, M., Srivastava, A., George, G.E., Cegan, J.C., Dettwiller, I.D., England, W.P., Farthing, M.W., Hunter, R.H., Lafferty, B., Linkov, I., Mayo, M.L., Parno, M.D., Rowland, M.A., Trump, B.D., Zhang-James, Y., Chen, S., Faraone, S.V., Hess, J., Morley, C.P., Salekin, A., Wang, D., Corsetti, S.M., Baer, T.M., Eisenberg, M.C., Falb, K., Huang, Y., Martin, E.T., McCauley, E., Myers, R.L., Schwarz, T., Sheldon, D., Gibson, G.C., Yu, R., Gao, L., Ma, Y., Wu, D., Yan, X., Jin, X., Wang, Y.-X., Chen, Y., Guo, L., Zhao, Y., Gu, Q., Chen, J., Wang, L., Xu, P., Zhang, W., Zou, D., Biegel, H., Lega, J., McConnell, S., Nagraj, V.P., Guertin, S.L., Hulme-Lowe, C., Turner, S.D., Shi, Y., Ban, X., Walraven, R., Hong, Q.-J., Kong, S., van de Walle, A., Turtle, J.A., Ben-Nun, M., Riley, S., Riley, P., Koyluoglu, U., DesRoches, D., Forli, P., Hamory, B., Kyriakides, C., Leis, H., Milliken, J., Moloney, M., Morgan, J., Nirgudkar, N., Ozcan, G., Piwonka, N., Ravi, M., Schrader, C., Shakhnovich, E., Siegel, D., Spatz, R., Stiefeling, C., Wilkinson, B., Wong, A., Cavany, S., España, G., Moore, S., Oidtman, R., Perkins, A., Kraus, D., Kraus, A., Gao, Z., Bian, J., Cao, W., Ferres, J.L., Li, C., Liu, T.-Y., Xie, X., Zhang, S., Zheng, S., Vespignani, A., Chinazzi, M., Davis, J.T., Mu, K., y Piontti, A.P., Xiong, X., Zheng, A., Baek, J., Farias, V., Georgescu, A., Levi, R., Sinha, D., Wilde, J., Perakis, G., Bennouna, M.A., Nze-Ndong, D., Singhvi, D., Spantidakis, I., Thayaparan, L., Tsiourvas, A., Sarker, A., Jadbabaie, A., Shah, D., Penna, N.D., Celi, L.A., Sundar, S., Wolfinger, R., Osthus, D., Castro, L., Fairchild, G., Michaud, I., Karlen, D., Kinsey, M., Mullany, L.C., Rainwater-Lovett, K., Shin, L., Tallaksen, K., Wilson, S., Lee, E.C., Dent, J., Grantz, K.H., Hill, A.L., Kaminsky, J., Kaminsky, K., Keegan, L.T., Lauer, S.A., Lemaitre, J.C., Lessler, J., Meredith, H.R., Perez-Saez, J., Shah, S., Smith, C.P., Truelove, S.A., Wills, J., Marshall, M., Gardner, L., Nixon, K., Burant, J.C., Wang, L., Gao, L., Gu, Z., Kim, M., Li, X., Wang, G., Wang, Y., Yu, S., Reiner, R.C., Barber, R., Gakidou, E., Hay, S.I., Lim, S., Murray, C., Pigott, D., Gurung, H.L., Baccam, P., Stage, S.A., Suchoski, B.T., Prakash, B.A., Adhikari, B., Cui, J., Rodríguez, A., Tabassum, A., Xie, J., Keskinocak, P., Asplund, J., Baxter, A., Oruc, B.E., Serban, N., Arik, S.O., Dusenberry, M., Epshteyn, A., Kanal, E., Le, L.T., Li, C.-L., Pfister, T., Sava, D., Sinha, R., Tsai, T., Yoder, N., Yoon, J., Zhang, L., Abbott, S., Bosse, N.I., Funk, S., Hellewell, J., Meakin, S.R., Sherratt, K., Zhou, M., Kalantari, R., Yamana, T.K., Pei, S., Shaman, J., Li, M.L., Bertsimas, D., Lami, O.S., Soni, S., Bouardi, H.T., Ayer, T., Adee, M., Chhatwal, J., Dalgic, O.O., Ladd, M.A., Linas, B.P., Mueller, P., Xiao, J., Wang, Y., Wang, Q., Xie, S., Zeng, D., Green, A., Bien, J., Brooks, L., Hu, A.J., Jahja, M., McDonald, D., Narasimhan, B., Politsch, C., Rajanala, S., Rumack, A., Simon, N., Tibshirani, R.J., Tibshirani, R., Ventura, V., Wasserman, L., O’Dea, E.B., Drake, J.M., Pagano, R., Tran, Q.T., Ho, L.S.T., Huynh, H., Walker, J.W., Slayton, R.B., Johansson, M.A., Biggerstaff, M., Reich, N.G.: Evaluation of individual and ensemble probabilistic forecasts of covid-19 mortality in the united states. Proceedings of the National Academy of Sciences 119(15), 2113561119 (2022). 10.1073/pnas.2113561119

[42] Mathis, S.M., Webber, A.E., León, T.M., Murray, E.L., Sun, M., White, L.A., Brooks, L.C., Green, A., Hu, A.J., Rosenfeld, R., Shemetov, D., Tibshirani, R.J., McDonald, D.J., Kandula, S., Pei, S., Yaari, R., Yamana, T.K., Shaman, J., Agarwal, P., Balusu, S., Gururajan, G., Kamarthi, H., Prakash, B.A., Raman, R., Zhao, Z., Rodríguez, A., Meiyappan, A., Omar, S., Baccam, P., Gurung, H.L., Suchoski, B.T., Stage, S.A., Ajelli, M., Kummer, A.G., Litvinova, M., Ventura, P.C., Wadsworth, S., Niemi, J., Carcelen, E., Hill, A.L., Loo, S.L., McKee, C.D., Sato, K., Smith, C., Truelove, S., Jung, S.-m., Lemaitre, J.C., Lessler, J., McAndrew, T., Ye, W., Bosse, N., Hlavacek, W.S., Lin, Y.T., Mallela, A., Gibson, G.C., Chen, Y., Lamm, S.M., Lee, J., Posner, R.G., Perofsky, A.C., Viboud, C., Clemente, L., Lu, F., Meyer, A.G., Santillana, M., Chinazzi, M., Davis, J.T., Mu, K., Pastore y Piontti, A., Vespignani, A., Xiong, X., Ben-Nun, M., Riley, P., Turtle, J., Hulme-Lowe, C., Jessa, S., Nagraj, V.P., Turner, S.D., Williams, D., Basu, A., Drake, J.M., Fox, S.J., Suez, E., Cojocaru, M.G., Thommes, E.W., Cramer, E.Y., Gerding, A., Stark, A., Ray, E.L., Reich, N.G., Shandross, L., Wattanachit, N., Wang, Y., Zorn, M.W., Aawar, M.A., Srivastava, A., Meyers, L.A., Adiga, A., Hurt, B., Kaur, G., Lewis, B.L., Marathe, M., Venkatramanan, S., Butler, P., Farabow, A., Ramakrishnan, N., Muralidhar, N., Reed, C., Biggerstaff, M., Borchering, R.K.: Evaluation of flusight influenza forecasting in the 2021–22 and 2022–23 seasons with a new target laboratory-confirmed influenza hospitalizations. Nature Communications 15(1), 6289 (2024). 10.1038/s41467-024-50601-9

[43] Tang, J., Rumack, A., Wilder, B., Rosenfeld, R.: Real-time forecasting of data revisions in epidemic surveillance streams. PLOS Computational Biology 21(11), 1–24 (2025). 10.1371/journal.pcbi.1013709

[44] White, L.A., León, T.M.: Forecastability of infectious disease time series: are some seasons and pathogens intrinsically more difficult to forecast? PLOS Computational Biology 22(4), 1–21 (2026). 10.1371/journal.pcbi.1014175

[45] Bracher, J., Ray, E.L., Gneiting, T., Reich, N.G.: Evaluating epidemic forecasts in an interval format. PLOS Computational Biology 17(2), 1–15 (2021). 10.1371/journal.pcbi.1008618

[46] Centers for Disease Control and Prevention: FluSight-forecast-hub. GitHub. Accessed: 2026-03-02. https://github.com/cdcepi/FluSight-forecast-hub

[47] Centers for Disease Control and Prevention: FluSight. Accessed: 2026-03-02. https://www.cdc.gov/flu-forecasting/index.html

[48] Chretien, J.-P., George, D., Shaman, J., Chitale, R.A., McKenzie, F.E.: Influenza forecasting in human populations: A scoping review. PLOS ONE 9(4), 1–8 (2014). 10.1371/journal.pone.0094130

[49] Hiller, K.M., Stoneking, L., Min, A., Rhodes, S.M.: Syndromic surveillance for influenza in the emergency department–a systematic review. PLOS ONE 8(9), 1–5 (2013). 10.1371/journal.pone.0073832

[50] Santillana, M., Nguyen, A.T., Dredze, M., Paul, M.J., Nsoesie, E.O., Brownstein, J.S.: Combining search, social media, and traditional data sources to improve influenza surveillance. PLOS Computational Biology 11(10), 1–15 (2015). 10.1371/journal.pcbi.1004513

[51] Hong, S., Son, W.-S., Park, B., Choi, B.Y.: Forecasting hospital visits due to influenza based on emergency department visits for fever: A feasibility study on emergency department-based syndromic surveillance. International Journal of Environmental Research and Public Health 19(19) (2022). 10.3390/ijerph191912954

[52] Bollerslev, T.: Generalized autoregressive conditional heteroskedasticity. Journal of Econometrics 31(3), 307–327 (1986). 10.1016/0304-4076(86)90063-1

[53] Keeling, M.J., Rohani, P.: Modeling Infectious Diseases in Humans and Animals. Princeton university press, Princeton, NJ, USA (2008)

[54] Center for Disease Control and Prevention: Yellow Book: Influenza (2025). https://www.cdc.gov/yellow-book/hcp/travel-associated-infections-diseases/influenza.html Accessed 2025-05-06

[55] Brauer, F., Castillo-Chavez, C., Feng, Z.: Models for influenza. In: Texts in Applied Mathematics. Texts in Applied Mathematics, pp. 311–350. Springer, New York, NY (2019)

[56] Goodman, J., Weare, J.: Ensemble samplers with affine invariance. Communications in Applied Mathematics and Computational Science 5(1), 65–80 (2010). 10.2140/camcos.2010.5.65

[57] Foreman-Mackey, D., Hogg, D.W., Lang, D., Goodman, J.: emcee: The MCMC Hammer. Publications of the Astronomical Society of the Pacific 125(925) (2013) https://arxiv.org/abs/1202.3665 [astro-ph.IM]. 10.1086/670067

[58] Howerton, E., Contamin, L., Mullany, L.C., Qin, M., Reich, N.G., Bents, S., Borchering, R.K., Jung, S.-m., Loo, S.L., Smith, C.P., Levander, J., Kerr, J., Espino, J., van Panhuis, W.G., Hochheiser, H., Galanti, M., Yamana, T., Pei, S., Shaman, J., Rainwater-Lovett, K., Kinsey, M., Tallaksen, K., Wilson, S., Shin, L., Lemaitre, J.C., Kaminsky, J., Hulse, J.D., Lee, E.C., McKee, C.D., Hill, A., Karlen, D., Chinazzi, M., Davis, J.T., Mu, K., Xiong, X., Pastore y Piontti, A., Vespignani, A., Rosenstrom, E.T., Ivy, J.S., Mayorga, M.E., Swann, J.L., España, G., Cavany, S., Moore, S., Perkins, A., Hladish, T., Pillai, A., Ben Toh, K., Longini, I., Chen, S., Paul, R., Janies, D., Thill, J.-C., Bouchnita, A., Bi, K., Lachmann, M., Fox, S.J., Meyers, L.A., Srivastava, A., Porebski, P., Venkatramanan, S., Adiga, A., Lewis, B., Klahn, B., Outten, J., Hurt, B., Chen, J., Mortveit, H., Wilson, A., Marathe, M., Hoops, S., Bhattacharya, P., Machi, D., Cadwell, B.L., Healy, J.M., Slayton, R.B., Johansson, M.A., Biggerstaff, M., Truelove, S., Runge, M.C., Shea, K., Viboud, C., Lessler, J.: Evaluation of the us covid-19 scenario modeling hub for informing pandemic response under uncertainty. Nature Communications 14(1), 7260 (2023). 10.1038/s41467-023-42680-x

[59] Stoner, O., Economou, T.: Multivariate hierarchical frameworks for modeling delayed reporting in count data. Biometrics 76(3), 789–798 (2019). 10.1111/biom.13188

[60] McGough, S.F., Johansson, M.A., Lipsitch, M., Menzies, N.A.: Nowcasting by bayesian smoothing: A flexible, generalizable model for real-time epidemic tracking. PLOS Computational Biology 16(4), 1–20 (2020). 10.1371/journal.pcbi.1007735

[61] Kline, D., Hyder, A., Liu, E., Rayo, M., Malloy, S., Root, E.: A bayesian spatiotemporal nowcasting model for public health decision-making and surveillance. American Journal of Epidemiology 191(6), 1107–1115 (2022). 10.1093/aje/kwac034

[62] Centers for Disease Control and Prevention National Health Safety Network: National Health Safety Network Hospital Respiratory Data. Accessed: 2026-03-02. https://data.cdc.gov/Public-Health-Surveillance/Weekly-Hospital-Respiratory-Data-HRD-Metrics-by-Ju/mpgq-jmmr/about_data

[63] United States Census Bureau: Census.gov, Business and Industry, Time series/Trend charts (2025). https://www.census.gov/econ/currentdata/?programCode=MARTS&startYear=1992&endYear=2025&categories[]=44W72&dataType=MPCSM&geoLevel=US&adjusted=0&notAdjusted=1&errorData=0#table-results Accessed 2025-05-13

[64] Biggerstaff, M., Cauchemez, S., Reed, C., Gambhir, M., Finelli, L.: Estimates of the reproduction number for seasonal, pandemic, and zoonotic influenza: a systematic review of the literature. BMC Infectious Diseases 14(1), 480 (2014). 10.1186/1471-2334-14-480

[65] Cheung, J.T.L., Tsang, T., Yen, H.-l., Perera, R.A.P.M., Mok, C.K.P., Lin, Y.P., Cowling, B., Peiris, M.: Determining existing human population immunity as part of assessing influenza pandemic risk. Emerging Infectious Disease journal 28(5), 977 (2022). 10.3201/eid2805.211965

[66] Gneiting, T., and, A.E.R.: Strictly proper scoring rules, prediction, and estimation. Journal of the American Statistical Association 102(477), 359–378 (2007). 10.1198/016214506000001437

[67] Centers for Disease Control and Prevention: ICD-10-CM. Accessed: 2026-04-15. https://www.cdc.gov/nchs/icd/icd-10-cm/index.html

[68] University of North Carolina School of Medicine: Respiratory Dashboard (2025). https://ncdetect.org/respiratory-dashboard/ Accessed 2025-05-06

[69] North Carolina Department of Health and Human Services: NC Respiratory Virus Summary Dashboard. Accessed: 2026-04-09. https://www.dph.ncdhhs.gov/programs/epidemiology/communicable-disease/respiratory-diseases/dashboard

[70] North Carolina Department of Health and Human Services: Detailed Respiratory Virus Surveillance Dashboard (2025). https://covid19.ncdhhs.gov/dashboard/respiratory-virus-surveillance Accessed 2025-05-06

[71] Center for Disease Control and Prevention: National, Regional, and State Level Outpatient Illness and Viral Surveillance (2025). https://gis.cdc.gov/grasp/fluview/fluportaldashboard.html Accessed 2025-05-06

[72] Raiffa, H., Schlaifer, R.: Applied Statistical Decision Theory. Student Edition. The M.I.T. Press, Cambridge, MA., USA (1961)

[73] Fink, D.: A Compendium of Conjugate Priors. Accessed: 2026-01-12. http://courses.physics.ucsd.edu/2018/Fall/physics210b/REFERENCES/conjugate_priors.pdf

[74] Gelman, A., Carlin, J.B., Stern, H.S., Dunson, D.B., Vehtari, A., Rubin, D.B.: Bayesian Data Analysis (3rd Ed.). Chapman and Hall/CRC, New York, USA (2013). 10.1201/b16018

